# Model-informed optimal allocation of limited resources to mitigate infectious disease outbreaks in societies at war

**DOI:** 10.1101/2024.08.01.24311365

**Authors:** Vaibhava Srivastava, Drik Sarkar, Claus Kadelka

## Abstract

Infectious diseases thrive in war-torn societies. The recent sharp increase in human conflict and war thus requires the development of disease mitigation tools that account for the specifics of war, such as scarcity of important public health resources. Differential equation-based compartmental models constitute the standard tool for forecasting disease dynamics and evaluating intervention strategies. We developed a compartmental disease model that considers key social, war, and disease mechanisms, such as gender homophily and the replacement of soldiers. This model enables the identification of optimal allocation strategies that, given limited resources required for treating infected individuals, minimize disease burden, assessed by total mortality and final epidemic size. A comprehensive model analysis reveals that the level of resource scarcity fundamentally affects the optimal allocation. Desynchronization of the epidemic peaks among several population subgroups emerges as a desirable principle since it reduces disease spread between different sub-groups. Further, the level of preferential mixing among people of the same gender, gender homophily, proves to strongly affect disease dynamics and optimal treatment allocation strategies, highlighting the importance of accurately accounting for heterogeneous mixing patterns. Altogether, the findings help answer a timely question: how can infectious diseases be best controlled in societies at war? The developed model can be easily extended to specific diseases, countries, and interventions.

**Significance statement:** Societies at war are particularly affected by infectious disease outbreaks, necessitating the development of mathematical models tailored to the intricacies of war and disease dynamics as valuable tools for policy-makers. The frequently limited availability of public health resources, such as drugs or medical personnel, yields a fundamental optimal allocation problem. This study frames this problem in a generic, modifiable context and proposes model-informed solutions by identifying allocation strategies that minimize disease burden, measured by total deaths or infections. The desynchronization of epidemic peaks among a heterogeneous population emerges as a general disease mitigation strategy. Moreover, the level of contact heterogeneity proves to substantially affect disease spread and optimal control.

## Introduction

In recent years, humankind has faced a surge in deadly infectious disease outbreaks, prompting remarkable global responses (*1*). Governments and international communities have shown unprecedented unity in implementing containment measures and managing these crises (*2, 3*). At the same time, armed conflict and war have also increased at an alarming rate (*4*). Disease control efforts become much more difficult when a country is engulfed in war or geopolitical conflict for a variety of reasons. Infectious diseases spread more rapidly, e.g., due to the large-scale movement of people, overcrowding in civilian shelters, poor sanitation, and generally deteriorating healthcare infrastructure, leading, among others, to interruptions in disease surveillance and control programs (*5, 6*).

One notable example is the 1918 influenza pandemic, which peaked in the final year of World War I. It infected an estimated one-third of the world population, with a global estimated death toll between 50 to 100 million people (*7*). The widespread movement of troops and displaced populations during World War I facilitated virus spread across national borders and even continents, often surpassing the capacity of local and national healthcare systems (*8*). A similar situation arose in Ukraine as a result of the ongoing war with Russia: SARS-CoV-2 and HIV-1 transmissions increased, particularly in areas where the healthcare infrastructure was severely compromised (*9, 10*). Even more recently, outbreaks of several different infectious diseases have been reported at camps for internally displaced people in Gaza, as well as among Israeli soldiers returning home from war (*11*). Other notable case studies include malaria outbreaks during social strife in Tajikistan in the mid 1990s (*12*), ongoing Ebola and pneumonic plague outbreaks in the war-torn Democratic Republic of Congo (*13–15*), a tularemia outbreak due to unsanitary environmental conditions following the Kosovo war in 1999-2000 (*16*), as well as several polio and yellow fever outbreaks in multiple African countries affected by war and forced migration (*6*).

Mathematical models have been an essential tool in the study of infectious diseases, providing critical insights into their spread and control. Among these, compartmental models, such as the classical SIR (Susceptible-Infected-Recovered) model, developed by Kermack and McKendrick nearly a hundred years ago, have proven historically effective (*17*).

Several mathematical models and innovative frameworks have been employed to better understand and mitigate the effects of conflict on disease transmission and control in regions affected by civil unrest and violence. For example, an extended SIR model that accounted for contaminated environments and hospital escapes due to violence revealed that ongoing conflict significantly worsened the tenth Ebola outbreak (2018-2020) in the Democratic Republic of the Congo (*18*). A recently developed framework conceptualized twelve impacts of war on social and environmental determinants of health and public health (*19*). Conflict-induced changes in human behavior and mobility can fundamentally affect the way infectious diseases spread throughout society at war. Therefore, interventions that proved effective or even optimal in traditional models may fall short when implemented during a conflict. This underscores the need for policymakers to develop comprehensive strategies that address the complexities of war-affected regions, ensuring that public health interventions remain robust and adaptable amid extraordinary challenges. Effective responses must encompass both targeted disease-specific interventions and broader health initiatives. However, there is a significant gap in generalizing these ideas to enable policymakers to optimally allocate public health resources, which are frequently severely limited, to mitigate infectious disease outbreaks in war-torn societies.

This manuscript partially fills this gap by answering the question: how should limited resources (e.g., drugs or healthcare personnel) required to treat people infected with a generic infectious disease be optimally allocated in a society at war? A standard, compartmental SIRD (Susceptible-Infected-Recovered-Deceased) model describes the spread of the disease (*20*) and enables us to derive general principles of optimal resource allocation. Standard SIR models make the default assumption that all individuals mix homogeneously. To capture the effect of war on contact structures and subsequent disease spread, the population is stratified by gender and location. A proportion of men, which are continuously replaced, fight at the war front where they may face higher disease transmission and case fatality rates (*21, 22*). A specific focus is on the concept of homophily, which describes the phenomenon that people with similar characteristics (in this work, gender) are more likely to interact. This is well-established in the social sciences (*23, 24*), but just recently began to gain traction in infectious disease models (*25–27*).

The model can be easily extended to account for additional heterogeneities that might affect the spread of the disease, such as age structure (*28, 29*) or spatial factors (*30, 31*). While the focus in this work is on the treatment of infected individuals, the model can also be modified to help policymakers in war-affected societies optimize a number of other public health interventions such as vaccination (*32*) or disease surveillance (*33*).

## Results and Discussion

We designed an infectious disease model that enables the study of public health resource allocation trade-offs in a society affected by war. Standard “SIR-type” differential equations describe how susceptible (S) individuals become infected/infectious (I) and how infected individuals eventually recover (R) or die (D). Beyond disease status, we stratified the population by gender (using, for simplicity, a binary classification into male and female) and by location (home (i.e., not-at-war) and war), making the simplifying assumption that all soldiers are male. We thus considered a total of 12 compartments: *X*_F_ representing females (at home), *X*_HM_ representing males at home, and *X*_WM_ representing males at war, where *X* ∈ {*S, I, R, D*}. We further assumed a fast-spreading pathogen such as SARS-CoV-2, allowing us to neglect births and deaths and changes in the population due to, e.g., migration. The pathogen can spread between the home and the spatially separated war front through the continuous replacement of soldiers.

The model further accounts for potential gender homophily, i.e., the fact that contacts at home may occur more frequently among people of the same gender. This is implemented using the homophily parameter *h* ∈ [0, 1]. Homogeneous mixing (i.e., no gender homophily) is modeled by *h* = 0, while *h* = 1 describes complete gender segregation. While hard to measure, the true level of gender homophily most likely lies somewhere between these two extremes (*23*).

Lastly, we assumed that infected individuals can be treated. Treatment shortens the time to recovery, yielding two related effects: (i) it lowers the probability that an infected individual dies from the disease, and (ii) it reduces the chance that an infected individual infects others. To capture the shortage of public health resources (doctors, nurses, drugs, etc.) during times of war, we assumed that the total capacity *c* ≥ 0 of these resources must be split between the home and the war front, with *τ* ∈ [0, 1] describing the proportion of resources allocated for the population at home. The effective treatment rates at home and at the war front decrease if treatment demand exceeds the allocated supply, causing slower recoveries, and more deaths and infections. All model parameters and their base values are described in Table 1.

**Table 1:** Description of model parameters.

| Parameter | Description | Base Value | Range |
| --- | --- | --- | --- |
| $\beta_H$ | Transmission rate at home | 0.6 | $[0, \infty)$ |
| $\beta_W$ | Transmission rate at war front | 0.8 | $[0, \infty)$ |
| $h$ | Gender homophily | 0 | $[0, 1]$ |
| $\eta$ | Soldier replacement rate | 0.05 | $[0, \infty)$ |
| $\kappa$ | Scaling factor for the severity of war | 1 | $[0, \infty)$ |
| $\gamma_H$ | Recovery rate at home | 0.2 | $[0, \infty)$ |
| $\gamma_W$ | Recovery rate at the war front | 0.2 | $[0, \infty)$ |
| $CFR_H$ | Case fatality rate at home | 0.05 | $[0, \infty)$ |
| $CFR_W$ | Case fatality rate at war front | 0.05 | $[0, \infty)$ |
| $\mu$ | Treatment rate | 0.2 | $[0, \infty)$ |
| $\phi_H$ | Treatment efficiency at home | varies | $[0, 1]$ |
| $\phi_W$ | Treatment efficiency at war | varies | $[0, 1]$ |

In what follows, we explore solutions to the public health decision-making problem: Given a total resource capacity *c*, find the optimal allocation strategy *τ*^*^(*c*) that minimizes disease burden, which can be assessed by various metrics, e.g., the overall number of disease-induced deaths, final epidemic size, or the basic and effective reproduction number. To begin, we focus on the latter.

### Optimal resource allocation to minimize the effective reproduction number

The basic reproduction number, denoted ℛ_0_, describes the expected number of secondary infections caused by one infected individual assuming the entire population is susceptible. The model was designed so that ℛ_0_ is invariant to changes in the assumed level of gender homophily, the gender distribution in the society (we later study a scenario where a disproportionate number of females have fled), and the proportion of people at war (Eq. 18). This enables an unconfounded identification of the effect of changes in these parameters on the optimal allocation of limited treatment resources.

While the two locations (home and war front) are presumed to be spatially separated, they are epidemiologically connected through the ongoing replacement of soldiers. ℛ_0,home_(*τ*) and ℛ_0,war_(*τ*) describe the basic reproduction number at home and at war, respectively (Eq. 19). For any choice of positive parameters, *∂*ℛ_0,home_(*τ*)*/∂τ* ≤ 0 and *∂*ℛ_0,war_(*τ*)*/∂τ* ≥ 0. As expected, allocating all treatment resources at home (*τ* = 1) thus minimizes the disease burden for the population at home (measured by ℛ_0,home_) while solely focusing on the war front (*τ* = 0) minimizes ℛ_0,war_.

While initially small, the number of infected increases as long as ℛ_0_(*τ*) = max*{*ℛ_0,home_(*τ*), ℛ_0,war_(*τ*)*} >* 1. Eventually, treatment demand may exceed available treatment resources, at which point infected individuals receive sub-optimal care, increasing their time to recovery and chance of dying from the disease (Eq. 3). The effective reproduction number *R*_eff_(*t*) describes the expected number of secondary infections caused by an infected at time *t* (Eq. 20). To illustrate how the optimal (i.e., *R*_eff_(*t*)-minimizing) treatment resource allocation *τ*^*^ depends on the amount of available resources *c*, we consider the following scenario: the transmission rate at war is 1*/*3 higher than at home (*β*_*H*_ = 0.3, *β*_*W*_ = 0.4), while the recovery rates are the same (*γ*_*H*_ = *γ*_*W*_ = 0.2) and equal to the treatment rate (*µ* = 0.2), implying that optimal treatment can cut the reproduction numbers in half. Moreover, *p*_SH_ = *p*_SW_ = 99% of females and males at home and at war are susceptible, while 1% is infected, and 25% of the total population (50% of males) are at the war front (*κ* = 1 and *p*_*M*_ = 0.5). If no treatment resources are available (*c* = 0), the effective reproduction rates at home and at war are *R*_eff, home_(*τ*) ≡ 1.5 and *R*_eff, war_(*τ*) ≡ 2, respectively (Fig. 1A). If very limited resources become available (e.g., *c* = 0.0005 as in Fig. 1B), it is, therefore, optimal to dedicate these resources entirely to the war front. As more treatment resources become available (e.g., *c* = 0.002 as in Fig. 1C), the optimal resource allocation *τ*^*^ shifts more and more towards a more balanced allocation between the two locations. At a resource capacity of about *c* = 0.006, all infected individuals can be optimally treated at a rate of *µ* if *τ*^*^ = 57% of all treatment resources are at home (Fig. 1D). Due to the assumed higher transmission rate at war, it proved optimal to allocate more resources per capita to the war front (note that 75% of the population is at home). If even more resources are available, a range of allocation values becomes, in theory, optimal (Fig. 1E). In practice, the allocation that puts the most emphasis on minimizing both ℛ_eff, home_ and ℛ_eff, war_ instead of only ℛ_eff_ should be preferable. In Fig. 1E, this implies the right endpoint of the interval of “optimal” allocation strategies is most preferable.

**Figure 1:**
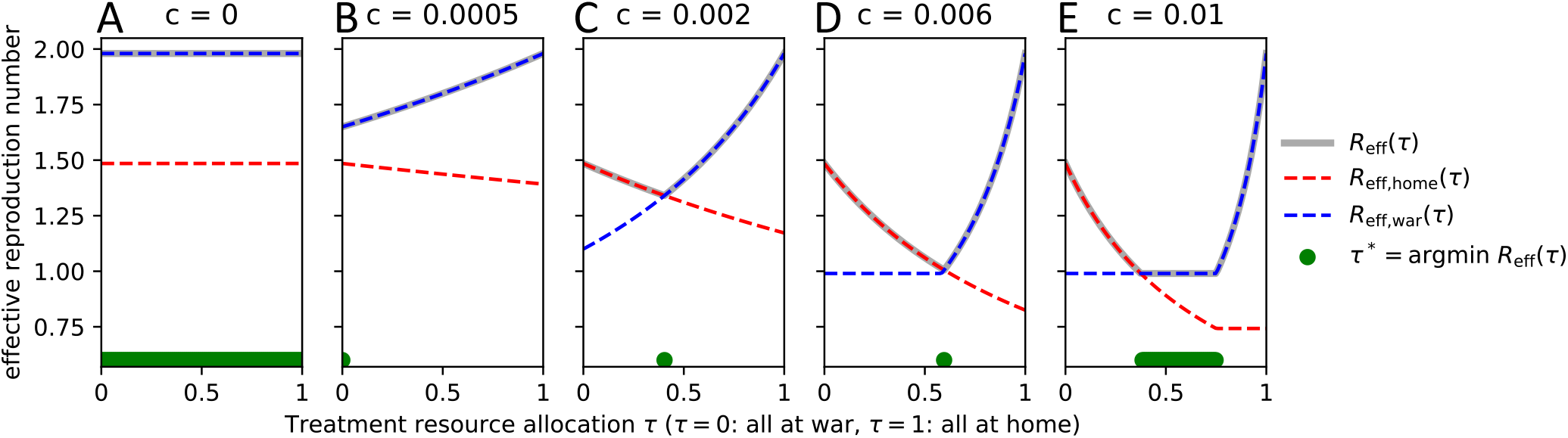
Dependence of the effective reproduction number on the treatment resource allocation *τ* for different levels of resource capacity *c*. (A) If no resources are available, *τ*does not affect the effective reproduction number, ℛ_eff_(*τ*). (B) As very limited resources become available, the initial focus is on the war front (*τ*^*^ = 0; green dots above the x-axis), (C) shifts more and more towards the home front (*∂τ*^*^*/∂c >* 0), (D) until at a resource capacity of about *c* = 0.006 perfect treatment of all infected individuals at both the home and the war front is possible (by allocating *τ*^*^ = 57% of resources to the home front). (E) At even higher resource capacities, many allocation strategies are optimal, as all treatment demand is met. The specific parameter choices here are *β*_*H*_ = 0.3, *β*_*W*_ = 0.4, *γ*_*H*_ = *γ*_*W*_ = 0.2, *µ* = 0.2, *κ* = 1 and 1% (99%) of each group *X* ∈ {F, HM, WM} were considered infected (susceptible). These choices explain why max_*c,τ*_ λ_2_ ≈ 1.5, max_*c,τ*_ λ_3_ ≈ 2, min_*c,τ*_ λ_2_ ≈ 0.75, min_*c,τ*_ λ_3_ ≈ 1.

### Optimal resource allocation to minimize total deaths and infections

Besides the basic and effective reproduction number, the final epidemic size (FES) and the total disease-induced mortality (i.e., the total number of infections and deaths that have occurred at the end of an epidemic) constitute two further important metrics when designing optimal policy decisions. While ℛ_eff_(*t*) and thus any ℛ_eff_(*t*)-minimizing treatment allocation strategy varies over time, total infections and deaths are computed once, at the end of the epidemic, making them better-suited for a resource allocation problems such as the one studied here. In fact, mortality is even the most frequently used metric in COVID-19 vaccine prioritization models (*34*).

If transmission rates at home and at the war front are comparable, the treatment allocation *τ* determines the size and the exact timing of the epidemic peaks as well as the number of deaths at home and at the war front (Fig. 2A-D). At the beginning of the epidemic (time *t* = 0), all infected individuals can be treated optimally. Once treatment demand in one location can no longer be sufficiently met, the effective reproduction number as well as the effective CFR increase, as seen, e.g., in Fig. 2A, E for infected at home at *t* = 10. This enables a positive feedback loop that yields more new infections and ultimately results in a high FES and death toll. As expected, the amount of resources allocated to one location correlated negatively with the location-specific number of infected and the death toll. The mortality-minimizing strategy *τ*^*^ = 79% (Fig. 2B, F) allocated treatment resources roughly according to the distribution of the population across the two locations (*κ* = 1 implies that 75% of people are home: all females, and half the males). The FES was minimized when a disproportionate amount of treatment resources were allocated at home (*τ*^*^ = 88%).

**Figure 2:**
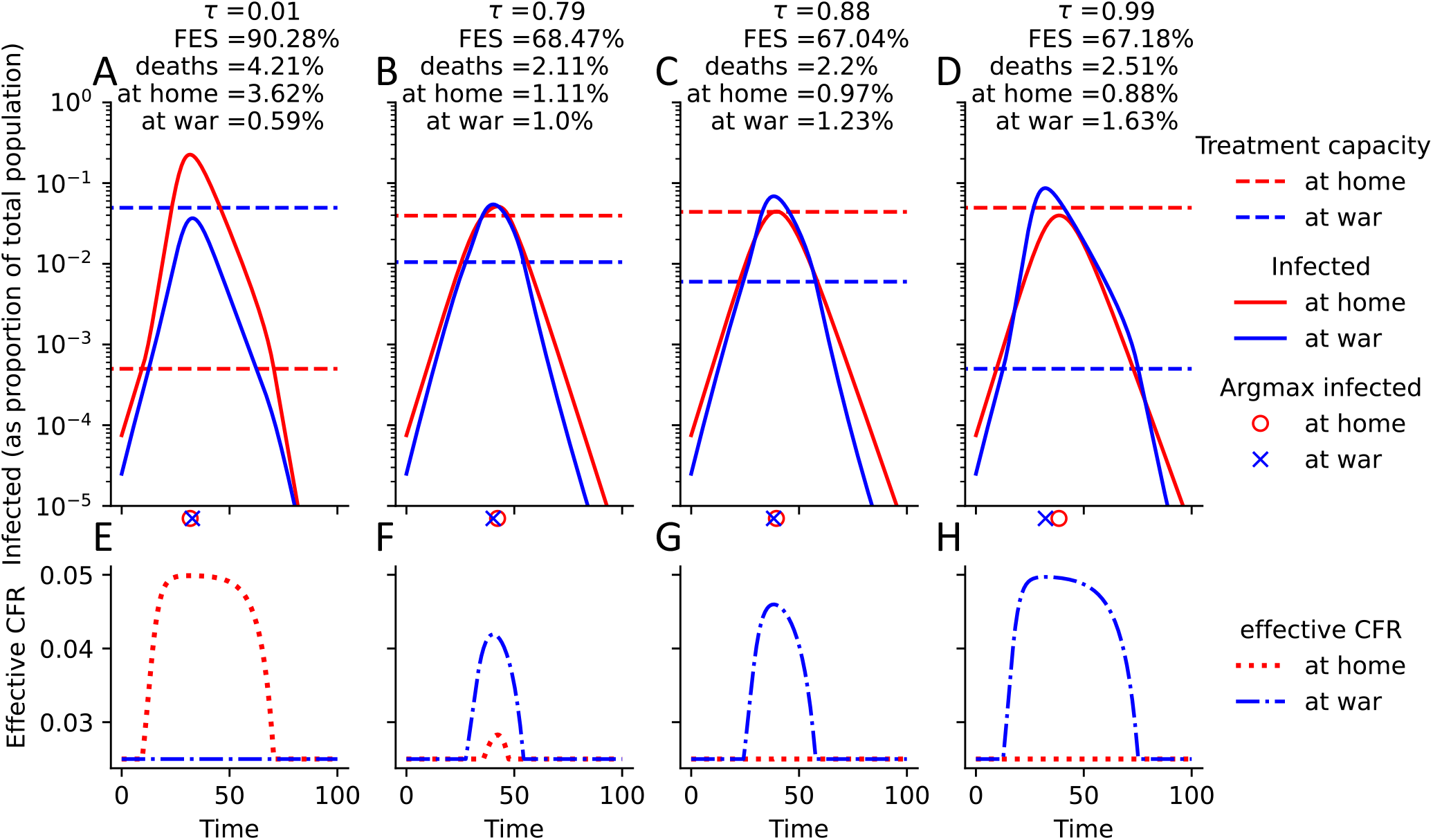
Comparison of epidemic curves when transmission rates at war and at home are the same. (A-D) For four different allocation strategies (A: *τ* = 0.01, B: *τ* = 0.79, which minimizes total deaths; C: *τ* = 0.88, which minimizes total infections; D: *τ* = 0.99), the location-stratified numbers of infected (red: home, blue: war) are shown over time. For each choice of *τ*, the total resource capacity is fixed at *c* = 0.05, and the location-specific, *τ*-dependent resource availabilities are indicated by dashed lines. The location-specific epidemic peak is indicated just below the x-axis. Final epidemic size (FES), total deaths, deaths at home and at war are shown at the top. All parameters are at their default value, as specified in Table 1, except for *β*_*W*_ = *β*_*H*_ = 0.6. (E-H) Corresponding plots of the effective case fatality rate (CFR), which is a function of the location-specific number of infected and treatment capacity, both shown in A-D.

In a situation where transmission rates at war are much higher than at home (e.g., due to crowded high-contact conditions or lack of hygiene), the epidemic progresses, as expected, much faster at the war front, irrespective of the treatment resource allocation (Fig. 3). The mortality-minimizing strategy in the specific scenario studied in Fig. 3 allocated 1 − *τ*^*^ = 80% of insufficient treatment resources to the war front, even though only 25% of the total population were assumed to be at the war front (Fig. 3B, F). Interestingly, this optimal resource allocation coincides with the point where optimal treatment of individuals at home can just be maintained. Allocating further resources to the war front leads to a much larger disease outbreak at home and thus more deaths, as seen in Fig. 3A, E. The final epidemic size was instead minimized by keeping *τ*^*^ = 66% of treatment resources at home (Fig. 3C, G). This choice strikes a balance between preventing a large outbreak at the more populous home front and reducing as many infections as possible at war.

**Figure 3:**
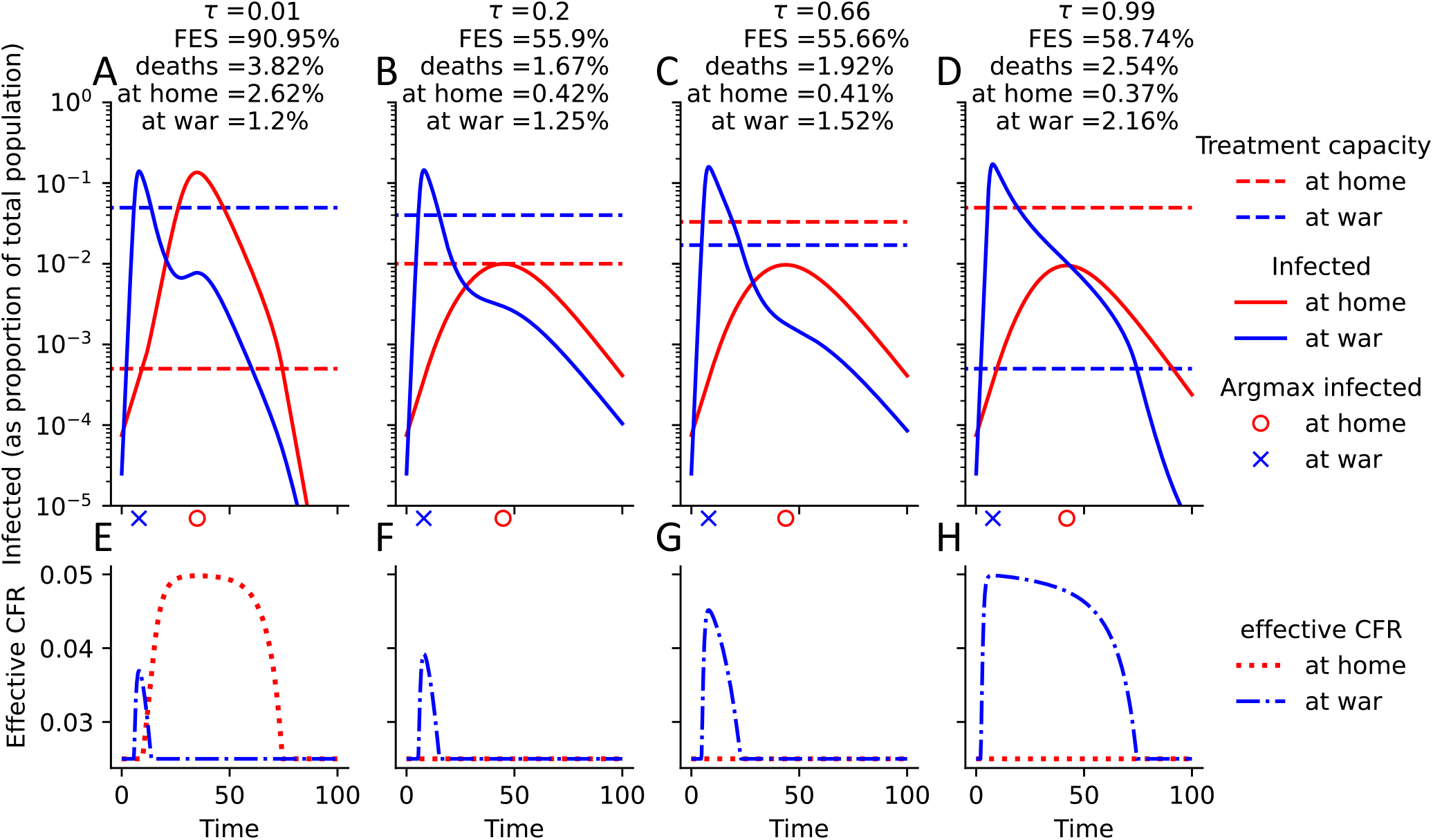
Comparison of epidemic curves when transmission rates at war are higher. (A-D) For four different allocation strategies (A: *τ* = 0.01, B: *τ* = 0.2, which minimizes total deaths; C: *τ* = 0.66, which minimizes total infections; D: *τ* = 0.99), the location-stratified numbers of infected (red: home, blue: war) are shown over time. For each choice of *τ*, the total resource capacity is fixed at *c* = 0.05, and the location-specific, *τ*-dependent resource availabilities are indicated by dashed lines. The location-specific epidemic peak is indicated just below the x-axis. Final epidemic size (FES), total deaths, deaths at home and at war are shown at the top. All parameters are at their default value, as specified in Table 1, except for *β*_*W*_ = 3*β*_*H*_ = 1.8. (E-H) Corresponding plots of the effective CFR (Eq. 3), which is a function of the location-specific number of infected and treatment capacity, both shown in A-D.

A comparison of Fig. 2 and Fig. 3 reveals an unexpectedly lower total death toll and FES when transmission rates at war are much higher. While higher transmission rates at war caused the expected higher number of infections and deaths at war, the desynchronization of the location-specific epidemic peaks implies that limited treatment capacities at home suffice to maintain the outbreak at a relatively modest size, which leads to fewer total infections and deaths (Fig. S1). During the strong early outbreak at the war front, males that move to the war front get infected at high rates and remain there while sick, thereby steadily deplenishing the pool of susceptible males at home. The desynchronized peaks are, therefore, a boon for the population at home. While we assumed treatment resource allocations to be fixed over time, such resources (e.g., doctors) can, in reality, be (at some rate) moved from one location to another based on needs. While not modeled here, the availability of partially mobile treatment resources would further reduce the disease burden in the case of desynchronized peaks.

### Impact of treatment capacity and gender homophily on optimal resource allocations

As expected, an increase in available treatment resources always resulted in fewer total deaths and infections (Fig. 4). Considering again a scenario where transmission rates at war are much higher than at home, total deaths and the FES were minimized by allocating very limited available resources entirely at home. If males and females were assumed to mix homogeneously (*h* = 0), this strategy proved optimal up to the point where the total resource capacity sufficed to treat all individuals at home optimally (Fig. 4A, B). All additional resources available beyond this point were then allocated to the war front. In the extreme case of complete gender segregation (*h* = 1), the optimal allocation strategy varied a lot more as the total resource capacity increased (Fig. 4C, D). Beyond a very small resource capacity (*c* = 0.0002), the focus on the war front increased (to *τ* = 0.4 at *c* = 0.005), then it decreased (to *τ* = 0.9 at *c* = 0.025) before it increased again (to *τ* = 0.2 at *c* = 0.15). An analysis of the epidemic trajectories at these four switch points revealed again the importance of a desynchronization of the epidemic peaks (Figs. S2, S3, S4,S5). Particularly for a resource capacity of *c* = 0.005, the outbreaks among the male population at home and at war are desynchronized by allocating 1 − *τ*^*^ = 60% of treatment resources to the war front (Fig. S3). Given that the true level of gender homophily lies somewhere between the two considered extremes (*23*), this interesting pattern highlights (i) the importance of accounting for homophily in infectious disease models and (ii) the complicated dynamics that arise from more realistic heterogeneous mixing patterns.

**Figure 4:**
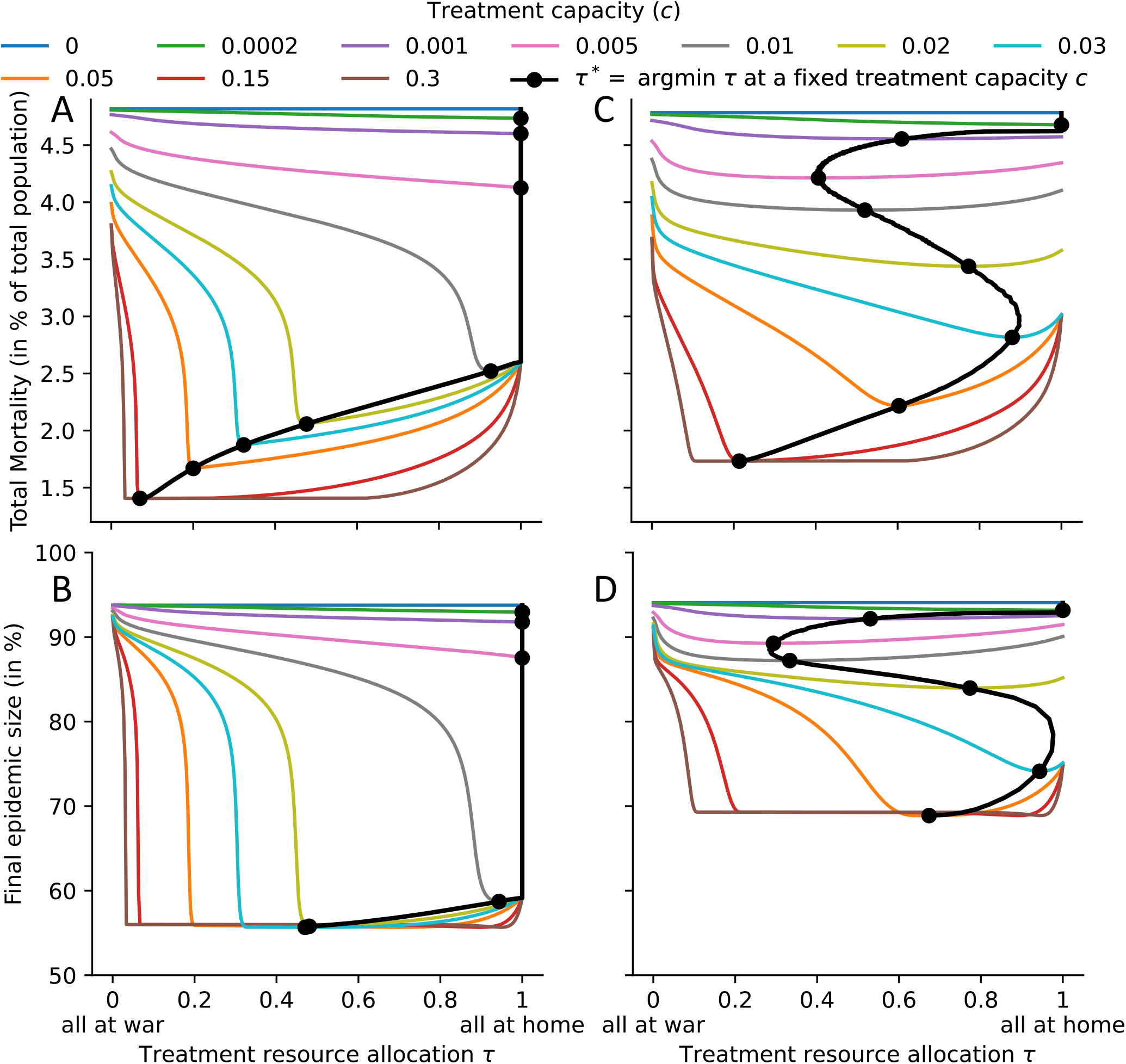
Model-predicted deaths and final epidemic size for various treatment capacities and allocation strategies. The black line indicates the optimal allocation strategy *τ*^*^ for any treatment capacity, with black dots indicating *τ*^*^ for the ten treatment capacities specified in the legend. The two extreme cases regarding gender homophily are considered: (A, B) no gender homophily (*h* = 0), (C, D) complete gender segregation (*h* = 1). Disease burden is quantified by (A, C) total mortality and (B, D) final epidemic size. All parameters are at their default value, as specified in Table 1, except for *β*_*W*_ = 3*β*_*H*_ = 1.8.

The FES proved much less affected by treatment inefficiencies due to insufficient resources during outbreaks. While total deaths decreased from close to 5% (the assumed CFR times the FES in the absence of any treatment) to roughly 1.4 − 1.7% (depending on the assumed level of gender homophily), the FES only decreased from about 95% to 57% in the case of no gender homophily, and even only to 70% in the case of complete gender segregation. Moreover, much fewer resources were required to achieve the maximal reduction in FES than the maximal reduction in total deaths. The lower impact of treatment on the FES can be explained by the fact that inefficient treatment causes more deaths in a direct and an indirect way: (i) the person receiving inefficient treatment faces a higher effective CFR (Eq. 3), and (ii) due to the longer time to recovery, this person also infects more people, which lowers treatment efficiencies further giving rise to the previously-mentioned positive feedback-loop and more deaths. Only the latter indirect effect contributes to higher final epidemic sizes if treatment is sub-optimal.

### Sensitivity of the optimal resource allocation

The model possesses a number of phenomenological parameters. While the effect of certain parameters on model outcomes is intuitively clear, others require further investigation. For example, increases in the CFR will result in corresponding increases in the total mortality without substantially affecting the FES (Fig. S6). On the other hand, an increased treatment rate will result in both fewer deaths and fewer infections (Fig. S7), and a slower soldier replacement rate will result in a weaker coupling between the disease dynamics at home and at the war front (Fig. S8). To assess the sensitivity of the optimal resource allocation to changes in less intuitive parameters, we varied the transmission rate at war (while keeping the transmission rate at home fixed), the severity of the war (scaled by the parameter *κ*, which determines the steady-state proportion of males at war), the outbreak severity (by modulating the reproduction number through the natural recovery rate), the level of gender homophily, as well as treatment capacity and allocation, as before. Since disease-induced mortality yielded thus far the most interesting results and is also the most frequently used metric in COVID-19 vaccine prioritization models (*34*), we focus from here only on minimizing total deaths.

Treatment, which was assumed to be fixed at a rate of *µ* = 0.2, was much more successful in reducing deaths if, assuming optimal treatment, ℛ_0,home_ = 1.5, ℛ_0,war_ ∈ [0.5, 4.5] (Fig. 5, rows 1 and 3) versus ℛ_0,war_ = 3, ℛ_0,home_ ∈ [1, 9] (Fig. 5, rows 2 and 4). The optimal treatment allocation strategy also varied more in the case of lower outbreak severity. In the case of very limited resources, a focus on the more populous home front minimized deaths unless transmission rates at war and at home were assumed to be similar (Fig. 5A-H). This finding was qualitatively consistent across changes in the outbreak severity, severity of war, and gender homophily. If ℛ_eff,home_ ≈ ℛ_eff,war_, the smaller size of the population at the war front implies that very limited treatment capacities can have a larger relative effect on reducing ℛ_eff,war_, thereby curbing infections and deaths more. At higher resource capacities, patterns already observed before emerged: If transmission rates at war are higher than at home and individuals mix homogeneously, it was optimal to focus on ensuring the best possible treatment of the large population at home before shifting attention to the war front (Fig. 5A, C), as seen in Fig. 4A, B. Due to the desynchronization of the epidemic peaks between the home and the war front, first described in Fig. 3, total deaths were lower despite substantially higher transmission rates at war (Fig. 5I, K). In the extreme case of complete gender segregation and higher at-war transmission rates (as already seen in Fig. 4C, D), the focus shifted, as treatment capacities increase, from home towards the war front, back to home and again back towards the war front, irrespective of the proportion of males at war (Fig. 5E, G). In case of higher transmission rates at home (unrealistic but worth exploring), it was optimal to use all treatment resources at home, irrespective of treatment capacity, gender homophily, outbreak severity, and gender homophily (Fig. 5A-H).

**Figure 5:**
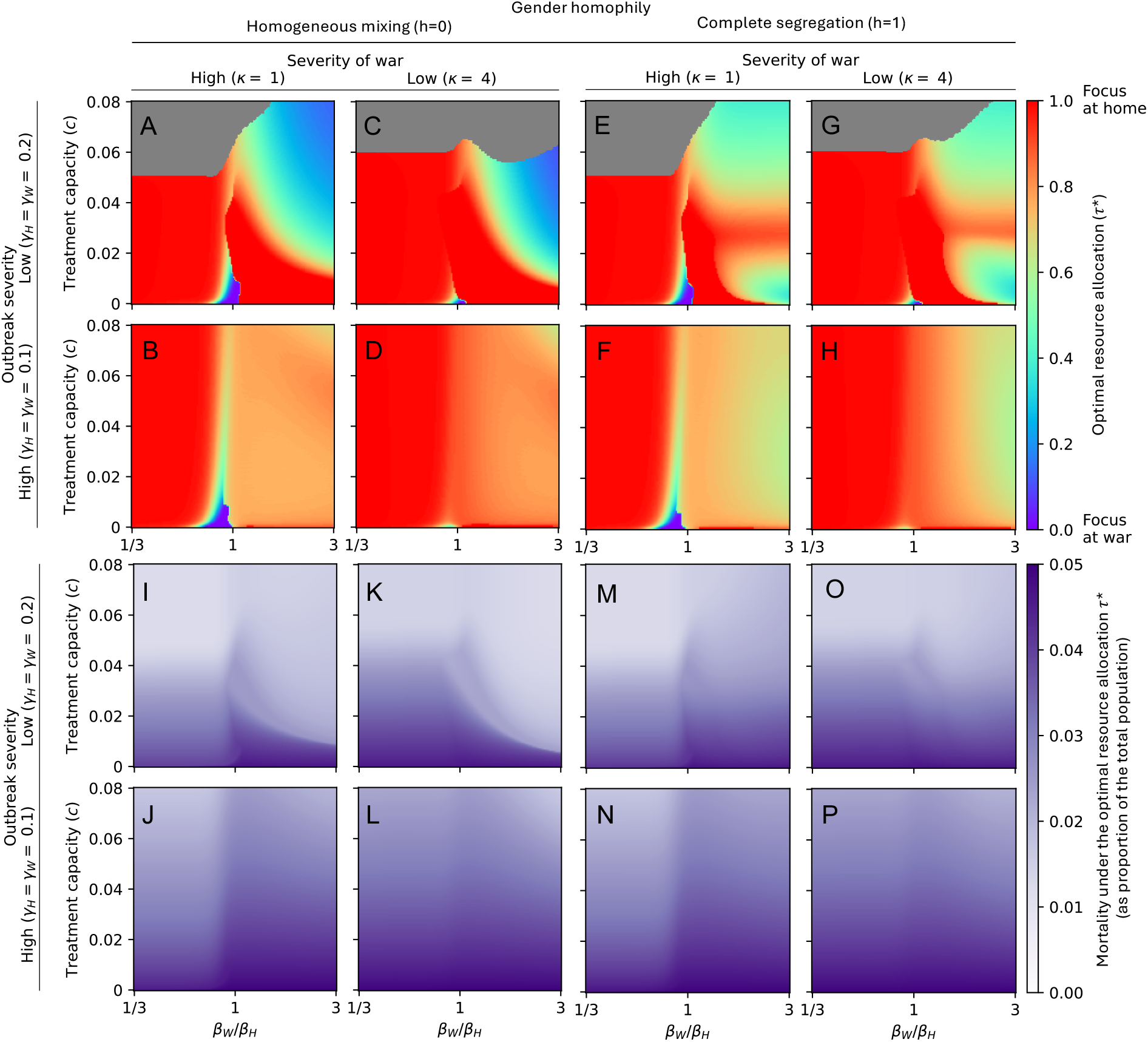
Sensitivity of the optimal resource allocation. For 150 equally-spaced values of treatment capacity (*c* ∈ [0, 0.08]) and transmission rates at war (*β*_*W*_ ∈ [1*/*3*β*_*H*_, 3*β*_*H*_]), the disease-induced mortality-minimizing treatment allocation strategy *τ*^*^ is shown in (A-H), while the total deaths under the respective strategy are shown in (I-P). The gray region describes parameter combinations where a range of resource allocations were optimal, i.e., where treatment resources sufficed to treat everyone optimally. In (A-D, I-L), males and females mix homogeneously (*h* = 0), while in (E-H, M-P), the extreme case of complete gender segregation (*h* = 1) is assumed. The outbreak severity varies between the rows, modulated by considering *γ*_*H*_ = *γ*_*W*_ = 0.2 (low severity) in rows 1 and 3, and *γ*_*H*_ = *γ*_*W*_ = 0.1 (high severity) in rows 2 and 4. The severity of war varies between the columns, modulated by considering *κ* = 1 (high severity, 50% of males at war) in rows 1 and 3, and *κ* = 4 (low severity, 20% of males at war) in rows 2 and 4. All other parameters are fixed at the default values described in Table 1.

### Impact of war-induced predominantly female migration

When war breaks out, men often stay, while women and children flee abroad, as exemplified by the overwhelmingly female wave of Ukrainian refugees since 2022 (*35*). This predominantly female migration due to war can give rise to a highly unbalanced gender distribution. When 70% (instead of 50%) of the population was assumed to be male, the focus on the war front increased for many parameter combinations, likely since fewer females at home required treatment (Fig. S9). In the case of lower transmission rates at war, the total mortality and the treatment capacity required to meet all demand both decreased slightly, likely because relatively more infections occurred at the war front. In general, however, the results proved qualitatively robust to changes in the male-to-female ratio.

## Conclusion

Conflict and war have risen substantially over the course of the last decade (*4*). Given the particular susceptibility of societies at war to infectious disease outbreaks (*6*), this emphasizes the need to study effective disease mitigation strategies. The frequent scarcity of public health resources in war-affected societies further complicates mitigation efforts, as decision-makers must choose between prioritizing the soldiers at war or the population at home. We framed this trade-off as a one-dimensional optimal resource allocation problem in a generic, extendable compartmental disease model and described the impact of several standard epidemiological parameters as well as parameters related to war dynamics and social structure.

The comprehensive model analysis revealed, among others, the desynchronization of epidemic peaks among subpopulations as a generally desirable principle that helps minimize disease burden, even when assuming fixed resource allocations. In reality, resources can be - at least slowly - reallocated, implying that strategies that spread out the epidemic peaks become even more beneficial. We only considered an initially synchronized outbreak (with the same initial disease incidence in each subgroup). Subsequent desynchronization is the result of differential transmission rates and treatment efficiencies due to the choice of allocation strategy. If the outbreak began in a specific subgroup, a different strategy might yield the desirable, maximal desynchronization.

A second major result is the importance of accounting for contact heterogeneity, exemplified in this study by the large impact of gender homophily on disease dynamics and optimal resource allocation strategies. In this study, we exclusively considered the extreme cases of homogeneous mixing and complete gender segregation. The true level of gender homophily, while hard to measure, will vary from country to country and lie somewhere between these extremes (*24*), leading to results that also are somewhere between the reported extreme results, as revealed in a one-dimensional sensitivity analysis (Fig. S10).

The developed model can be easily extended and adapted to specific pathogens and situations, yielding more accurate model dynamics at the expense of higher complexity. For example, incorporating a non-zero rate of female participation in warfare could capture more accurately current military trends (*36, 37*). Similarly, age-assortative mixing patterns have become a crucial building block of accurate infectious disease models (*38*). SIR dynamics fail to accurately describe the spread of certain pathogens, such as SARS-CoV-2, which is frequently transmitted prior to the onset of symptoms. The inclusion of additional compartments could mitigate this issue. A further model limitation relates to pathogens with substantial presymptomatic or asymptomatic transmission. We assumed that infected males do not move from and to the war front. This is only possible if cases can be ascertained, requiring either clear symptoms or efficient testing. Therefore, a likely more realistic approach would be to assume lower (but non-zero) replacement rates for infected individuals. Altogether, there exists a plethora of possible modifications to the developed model. We consciously employed a very simple, abstract model to facilitate the analysis and to focus on the key social and disease mechanisms that affect disease spread and optimal mitigation strategies in a population affected concurrently by war and an infectious disease outbreak.

## Material and Methods

### Compartmental disease model

We describe the spread of infectious disease among the population of a country at war using a system of twelve differential equations, each describing the size of one subpopulation (also referred to as a compartment). The population is divided into three different subgroups based on gender (females and males) and location (those at home and those at the spatially separated war front), and we make the simplifying assumption that only men fight at war. Distinguishing further four disease statuses yields the following twelve compartments: susceptible individuals (*S*_F_, *S*_HM_, *S*_WM_), infected individuals (*I*_F_, *I*_HM_, *I*_WM_), recovered individuals (*R*_F_, *R*_HM_, *R*_WM_), and deceased individuals (*D*_F_, *D*_HM_, *D*_WM_). The subscripts denote the gender and location (F for females, HM for males at home, WM for males at war). The total population size *N* is fixed when including deceased individuals in the count. Balancing model complexity and accuracy, we do not model births, deaths, and active migration since the generic infectious disease studied here is assumed to be fast-spreading with a short disease generation time. Note that the impact of migration on model predictions can be indirectly assessed by varying the initial conditions, i.e., the male-to-female ratio, as females and children often leave war-torn regions at higher rates (*35*).

#### Continuous replacement of soldiers

We assume that the home and the war front are spatially separated, implying that no contacts between men at war and the rest of the population occur. The disease dynamics at home and at war are nevertheless coupled due to the replacement of males at war with males at home, and vice versa, at fixed rates of *κη* and *η*, respectively. Here, *η* describes the speed of the replacement, while *κ* represents the severity of the war. We assume that infected individuals are not replaced until they recover. If *p*_*M*_ ∈ [0, 1] denotes the proportion of the population that is male; we have at any steady state

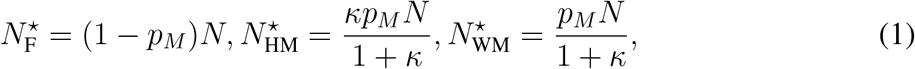

where 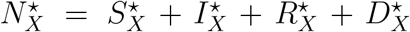 denotes the total population size of subgroup *X* ∈ {F, HM, WM} at a steady state.

#### New infections and gender homophily

The force of infection, FOI_*X*_, describes the rate at which susceptible individuals in a given subgroup *X* ∈ {F, HM, WM} become infected. It depends on the proportion of infected individuals, the contact patterns among subgroups, mitigation measures, as well as the location-specific transmission rates, *β*_*H*_ and *β*_*W*_. By considering 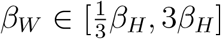, we study situations in which the transmission rates between the two locations differ up to a factor of three, due to e.g., more or prolonged contacts in one location. At home, we assume that females and males have the same total number of contacts. In the absence of preferential mixing among people of the same gender, we assume that the proportion of contacts an individual at home has with females is *N*_F_*/*(*N*_F_ + *N*_HM_) and with males is *N*_HM_*/*(*N*_F_ + *N*_HM_), irrespective of the gender of the individual itself (note that *N*_*X*_ = *N*_*X*_(*t*) denotes the total population size of subgroup *X* ∈ {F, HM, WM} and does typically not vary much over time). To account for possible gender homophily *h* ∈ [0, 1] (that is, more frequent contacts between people of the same gender due to preference and/or job setting), we move a fraction *h* of contacts among people of the opposite gender to contacts among people of the same gender, as explained in detail in (*39*). This approach ensures the same level of total contact between the population at home, irrespective of the assumed level of gender homophily.

Together, these considerations yield subgroup-specific forces of infection

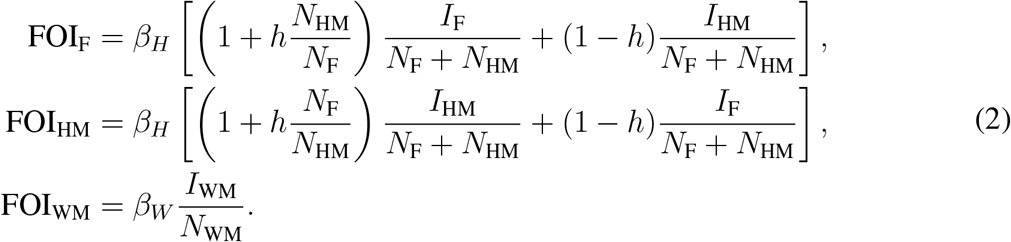

#### Natural recovery and death

Location-specific recovery rates (*γ*_*H*_ ≥ 0 for females and males at home, and *γ*_*W*_ ≥ 0 for males at war) and case fatality rates (*CFR*_*H*_ ≥ 0 for females and males at home, and *CFR*_*W*_ ≥ 0 for males at war) conclude the description of the disease dynamics. Upon leaving the infected compartment, any individual recovers with probability 1 − *CFR* and dies with probability *CFR*. While they may differ in practice, we only considered the situation where *γ*_*H*_ = *γ*_*H*_ and *CFR*_*H*_ = *CFR*_*W*_.

#### Treatment

We account for the possibility of treatment of infected individuals by including an additional recovery rate due to treatment, implying that treatment accelerates recovery. The treatment rate is *µϕ*_*H*_ for those at home and *µϕ*_*W*_ for those at war. Here, *µ* ≥ 0 describes the maximal possible treatment rate (given unlimited resources) and the location-specific treatment efficiencies *ϕ*_*H*_, *ϕ*_*W*_ ∈ [0, 1] enable the definition of an optimal control limited-resource allocation problem (see below). Individuals who recover due to treatment do not die. Treatment thus has two effects: (i) it shortens the time an individual is infected and can infect others, and (ii) it lowers the proportion of infected individuals that die from the disease.

To fully understand these two effects, it helps to consider a single infected individual at location *X* ∈ *{H, W}*. The individual’s time to recovery due to treatment and the time to natural recovery is exponentially distributed with parameters *µϕ*_*X*_ and *γ*_*X*_, respectively. When the individual is not treated, it dies with probability *CFR*_*X*_. Otherwise, it is assumed to survive. Thus, the probability that an infected dies from the disease depends on the treatment rate *µϕ*_*X*_, and is termed the effective CFR. Thus, given two independent random variables *A* ∼ Exp(*µϕ*_*X*_), *B* ∼ Exp(*γ*_*X*_) with joint density function *p*(*a, b*) = *µϕ*_*X*_*γ*_*X*_ exp(−*µϕ*_*X*_*a* − *γ*_*X*_*b*), we have effective CFR_*X*_ = CFR_*X*_ ℙ (*A > B*)

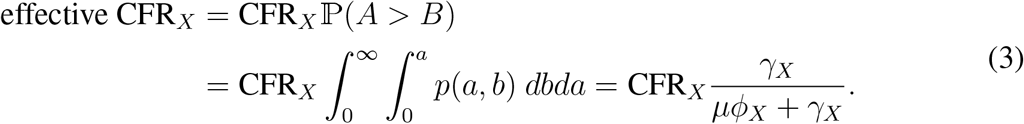

### Complete model and parameter choices

Transitions between the twelve compartments are illustrated in Fig. 6, and the complete set of model equations is as follows:

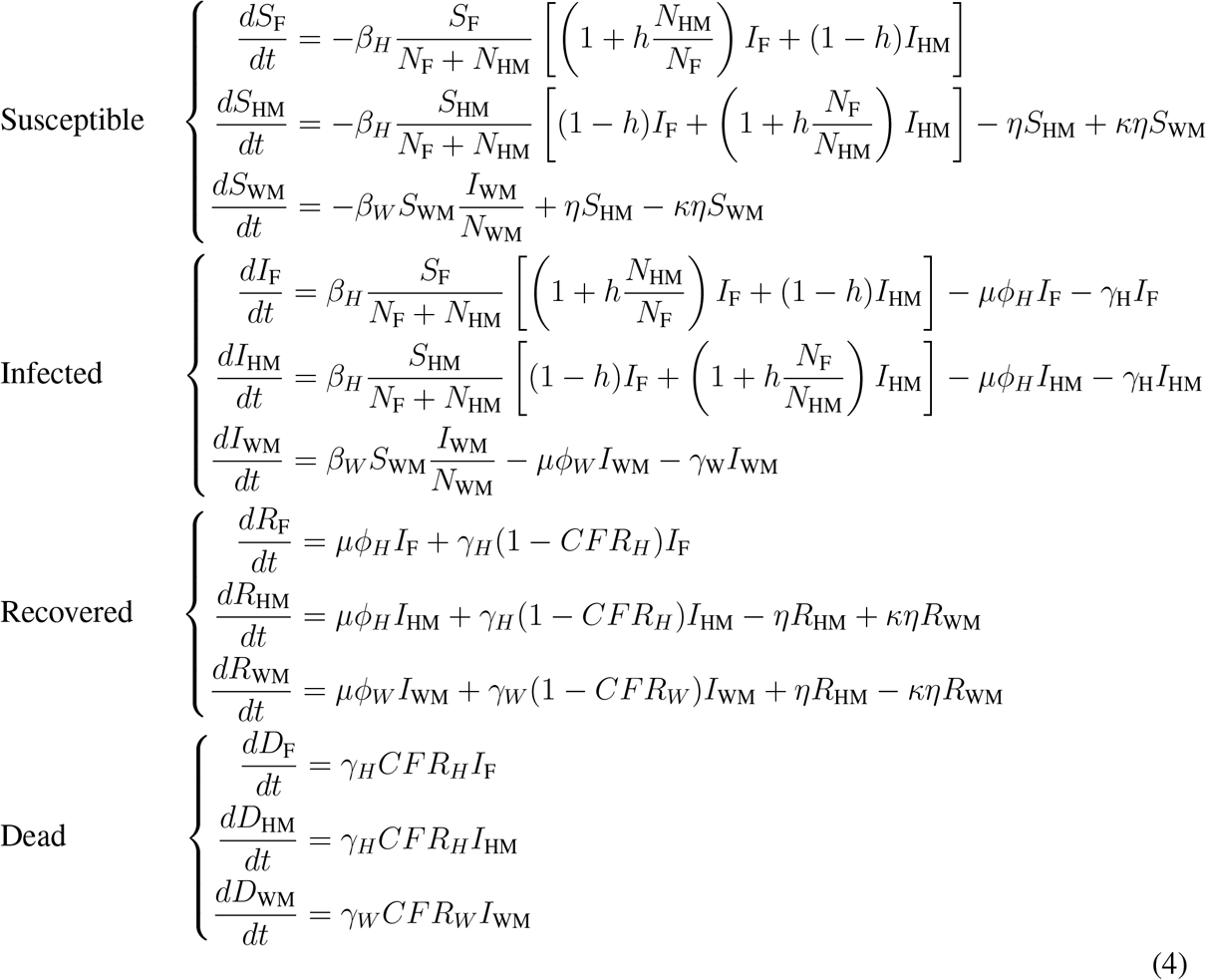

**Figure 6:**
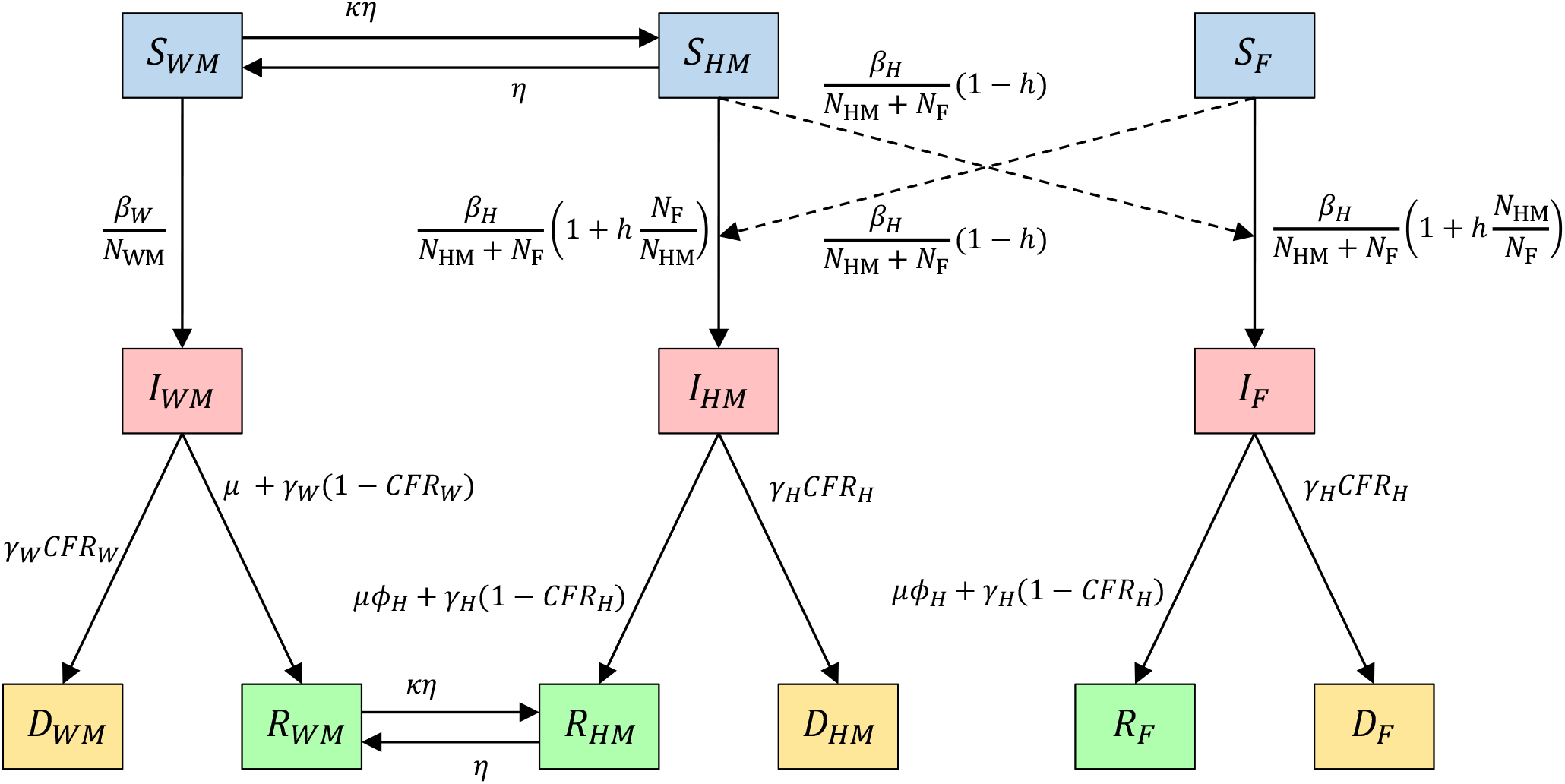
Illustration of the modeled transitions (Eq. 4) between the different compartments (susceptible: *S*_*X*_, infected: *I*_*X*_, recovered: *R*_*X*_, and dead: *D*_*X*_), where the subgroup *X* describes location (home: H, and war: W) and gender (male: M, and female: F). The parameters are defined in Table 1.

Given the abstract nature of the model, we set all parameters to default values (Table 1) and conducted extensive sensitivity analyses to explore the impact of specific parameters.

### Optimal control resource allocation problem

Countries at war often suffer from insufficient public health resources, hindering the effective control of an infectious disease outbreak. In this study, we consider a one-dimensional resource allocation problem: given *c* > 0 people can be treated at any moment (where *c* is determined, among others, by the number of available healthcare personnel and drugs), how should the treatment resources be optimally split between the war and the home front to minimize the overall number of disease-induced deaths.

Given the presumed spatial separation between the home and the war front and the fast outbreak dynamics, we considered the resource allocation fixed over time and let *τ* ∈ [0, 1] describe the proportion of resources allocated at home. Then, *τc* and (1 − *τ*)*c* represent the maximal number of infected individuals at home, and at war, respectively, that can be treated at the optimal rate *µ*. If more individuals are infected at either location, treatment becomes sub-optimal, and the effective treatment rate decreases proportionally. That is, the effective treatment rate at home and at war is *µϕ*_*H*_(*t*) and *µϕ*_*W*_ (*t*), where the treatment efficiencies are

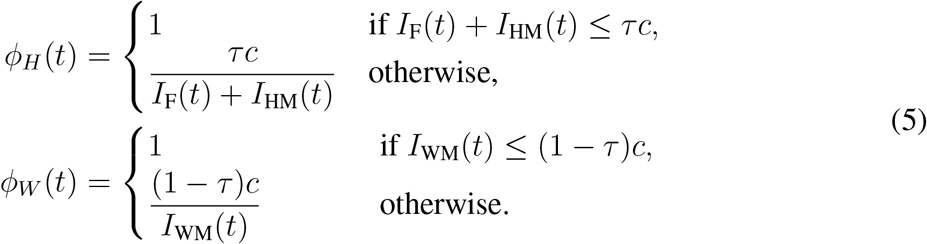

The optimal control problem thus is: Given a treatment capacity of *c* > 0, find the treatment resource allocation *τ*^⋆^(*c*) ∈ [0, 1] that minimizes the disease burden, quantified in this study by the overall mortality

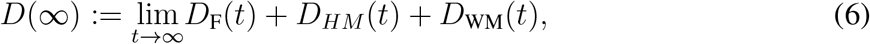

the final epidemic size (FES)

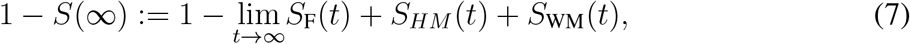

or the effective reproduction number at a certain moment in time, defined below.

### Simulations

To compute the total number of disease-induced deaths (Eq. 6) and the FES (Eq. 7) for a given choice of parameters and resource allocation *τ*, we solved the system of differential equations numerically using the fourth-order Runge-Kutta method (RK4) and a time step Δ*t* = 0.25. This method is accurate and computationally efficient. The use of the high-performance Python compiler Numba substantially improved the compute time (*40*).

Throughout, we initiated the disease dynamics at an early stage of the outbreak, assuming 0.01% of people are infected, while 99.99% are susceptible. The infected individuals are split across the three subgroups proportional to the size 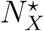 of each subgroup *X* ∈ {F, HM, WM} at the disease-free equilibrium (Eq. 1).

### Derivation of the basic reproduction number

The basic reproduction number, ℛ_0_, constitutes one of the most widely used metrics in the study of infectious diseases. It quantifies the average number of secondary infections caused by a single infected individual in a completely susceptible population. Following the standard next-generation matrix approach to compute ℛ_0_ (*41, 42*) and writing the compartments as

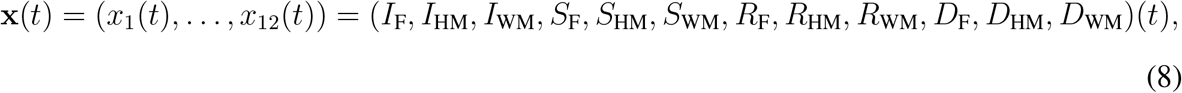

the disease-free equilibrium can be written as

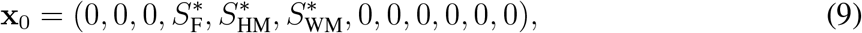

where 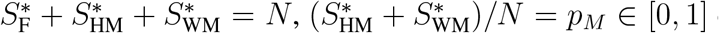 describes the proportion of the population that is male, and 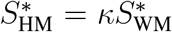 relates the number of males at home and at war. As in Eq. 1, for a given *p*_*M*_ we have

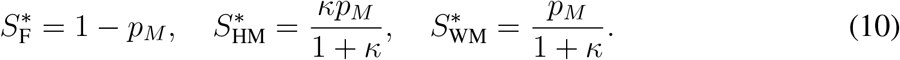

The disease dynamics of the model can then be represented as

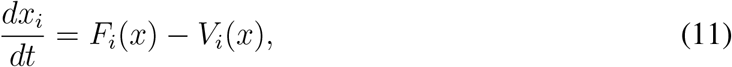

where *F*_*i*_(*x*) describes the appearance of newly infected individuals in compartment *i*, while *V*_*i*_(*x*) describes the rate of net out- versus in-flow of individuals in compartment *i*. We have

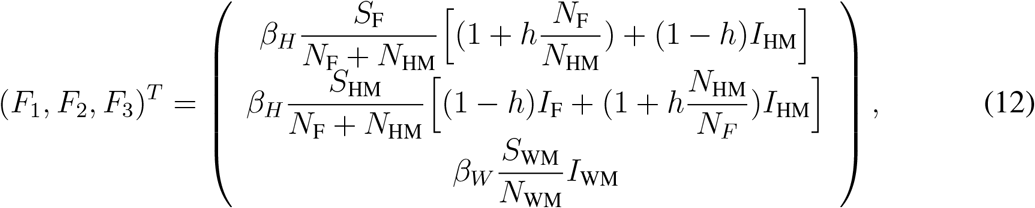

and

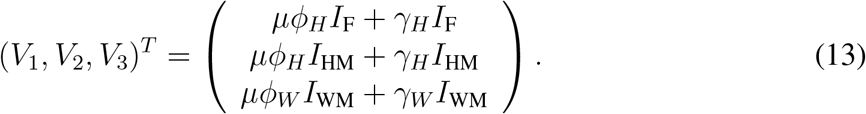

As described in (*42*), we can compute two Jacobian *m* × *m*-matrices *F* = *∂F*_*i*_*/∂x*_*j*_(**x**_0_) and *V* = *∂V*_*i*_*/∂x*_*j*_(**x**_0_) such that ℛ_0_ = *ρ*(*FV* ^−1^) where *ρ*(·) denotes the spectral radius of a square matrix. For the presented model, *FV* ^−1^ is a block matrix (since people at war and home cannot infect each other) with eigenvalues

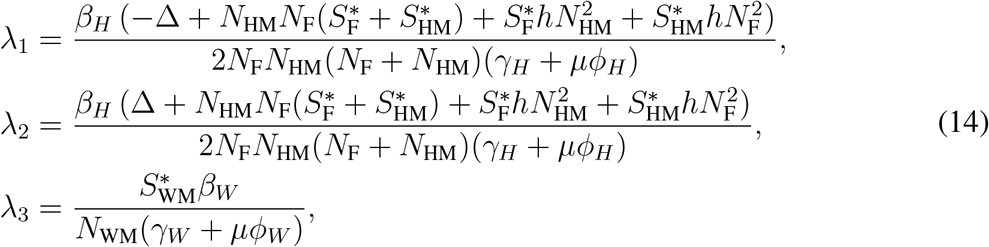

where

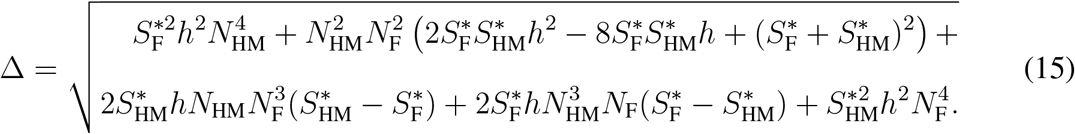

Since 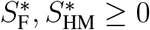 we have |λ_1_| *<* |λ_2_| such that the basic reproduction number ℛ_0_ is

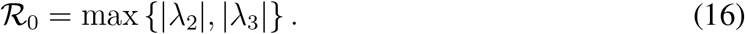

As long as some treatment resources are available (i.e., *c* > 0) and these resources are somewhat distributed between the home and war front (i.e., 0 < *τ* < 1), we have at the disease-free equilibrium, *ϕ*_*H*_ = *ϕ*_*W*_ = 1. That is, the initially tiny fraction of the population that is infected can be treated at the optimal rate *µ*. Moreover at the disease-free equilibrium, 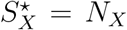 for *X* ∈ {F, HM, WM} so that

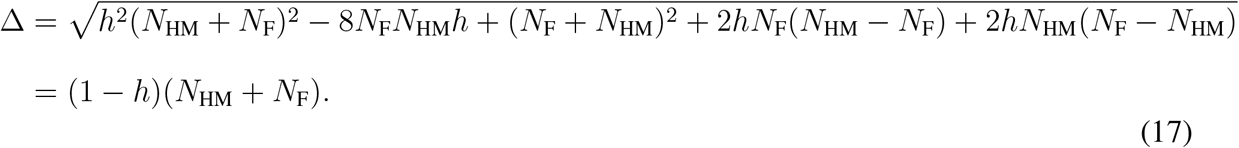

With this, Eq. 16 becomes

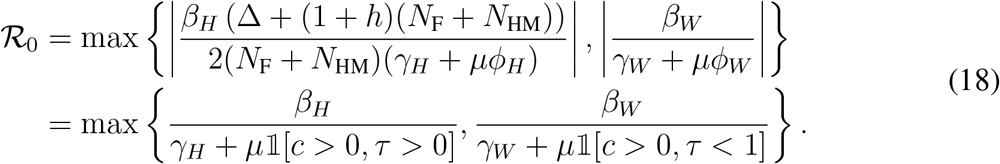

Since the two locations (home and war) are spatially separated, λ_2_(*τ*) describes the basic reproduction number at home, while λ_3_(*τ*) describes the basic reproduction number at the war front. That is,

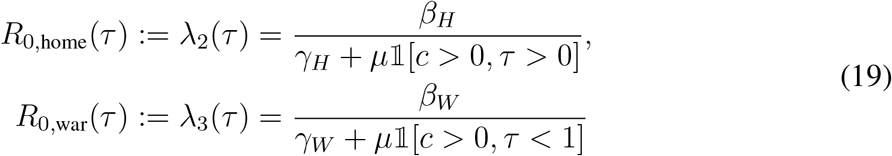

Eq. 18 further highlights that neither the relative number of individuals in the three groups, F, HM, WM, nor the level of gender homophily influences ℛ_0_. This validates the model setup and enables us to study the true impact of homophily and changes in the population distribution (through varying initial conditions) on optimal treatment resource allocation without having to worry about a confounding effect due to changes in the basic reproduction number.

### Derivation of the effective reproduction number

When more than a negligibly small proportion of the population is infected (some may even have recovered), the basic reproduction number no longer applies because it is defined as the number of secondary infections caused by one infected individual in a *fully susceptible* population. Instead, we use the *effective reproduction number*, denoted ℛ_eff_(*t*), to describe the expected number of secondary infections caused by one infected individual at time *t*. As long as the proportion of females and males at home that are susceptible is the same, denoted *p*_SH_(*t*) ∈ [0, 1], the expressions in Eq. 14 can be simplified, as shown above for ℛ_0_, to obtain

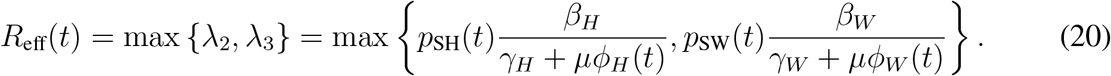

Here, *p*_SW_(*t*) ∈ [0, 1] describes the proportion of males at war that are still susceptible, and the treatment efficiencies *ϕ*_*H*_(*t*), *ϕ*_*W*_ (*t*) are functions of the location-specific number of infected, the resource allocation strategy *τ*and the total resource capacity *c*, and can therefore take on any value in [0, 1]. Again, as for ℛ_0_, λ_2_(*τ*) describes the effective reproduction number at home, while λ_3_(*τ*) describes the effective reproduction number at the war front. That is,

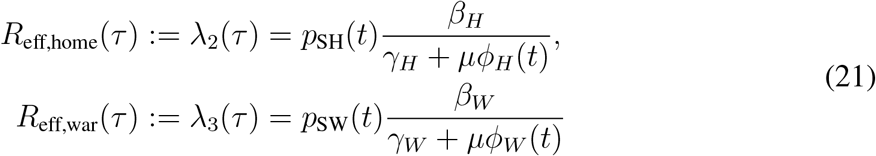

If the proportion of males and females at home that are susceptible is not the same, we can still compute *R*_eff_(*τ*), *R*_eff,home_(*τ*), and *R*_eff,war_(*τ*) from Eqs. 14,15 but cannot obtain formulas as simple as Eqs. 20,21.

## Data Availability

The Python code used to run the infectious disease model and generate all figures is provided at https://github.com/ckadelka/disease-control-at-war.

## Funding

VS gratefully acknowledges financial support from the Johnston-Peters Summer Graduate Assistantship. CK was partially supported by a travel grant from the Simons Foundation (grant number 712537).

## Author Contributions

Conceptualization: VS, DS, CK; Formal analysis: VS, CK; Methodology: VS, DS, CK; Software: VS, CK; Visualization: VS, CK; Supervision: CK; Writing: VS, DS, CK.

## Competing interests

The authors declare that they have no competing interests.

## Supplementary Materials

**Figure S1:**
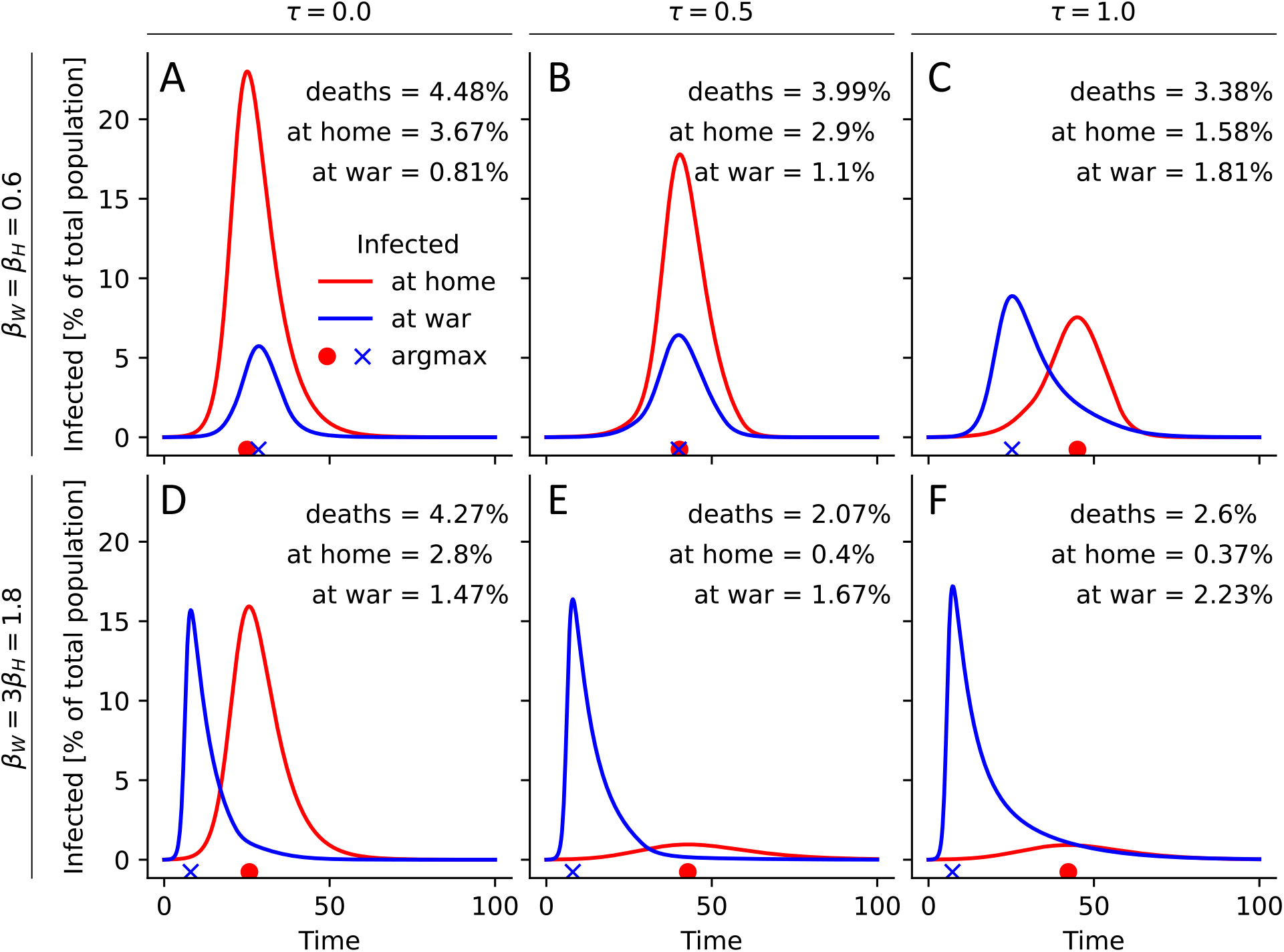
Comparison of epidemic curves for different relative transmission rates and treatment allocations. For three different treatment allocation strategies (A, D: *τ* = 0 (all at war), B, E: *τ* = 0.5, C, F: *τ* = 1 (all at home)), the location-stratified numbers of infected (red: home, blue: war) are shown over time. For each choice of *τ*, the total resource capacity is fixed at *c* = 0.05, and dashed lines indicate the location-specific, *τ*-dependent resource availabilities. The location-specific epidemic peak is indicated just below the x-axis. Total deaths, as well as location-specific deaths, are shown at the top. All parameters are at their default value, as specified in Table 1, except for (A-C) *β*_*W*_ = *β*_*H*_ = 0.6 and (D-F) *β*_*W*_ = 3*β*_*H*_ = 1.8.

**Figure S2:**
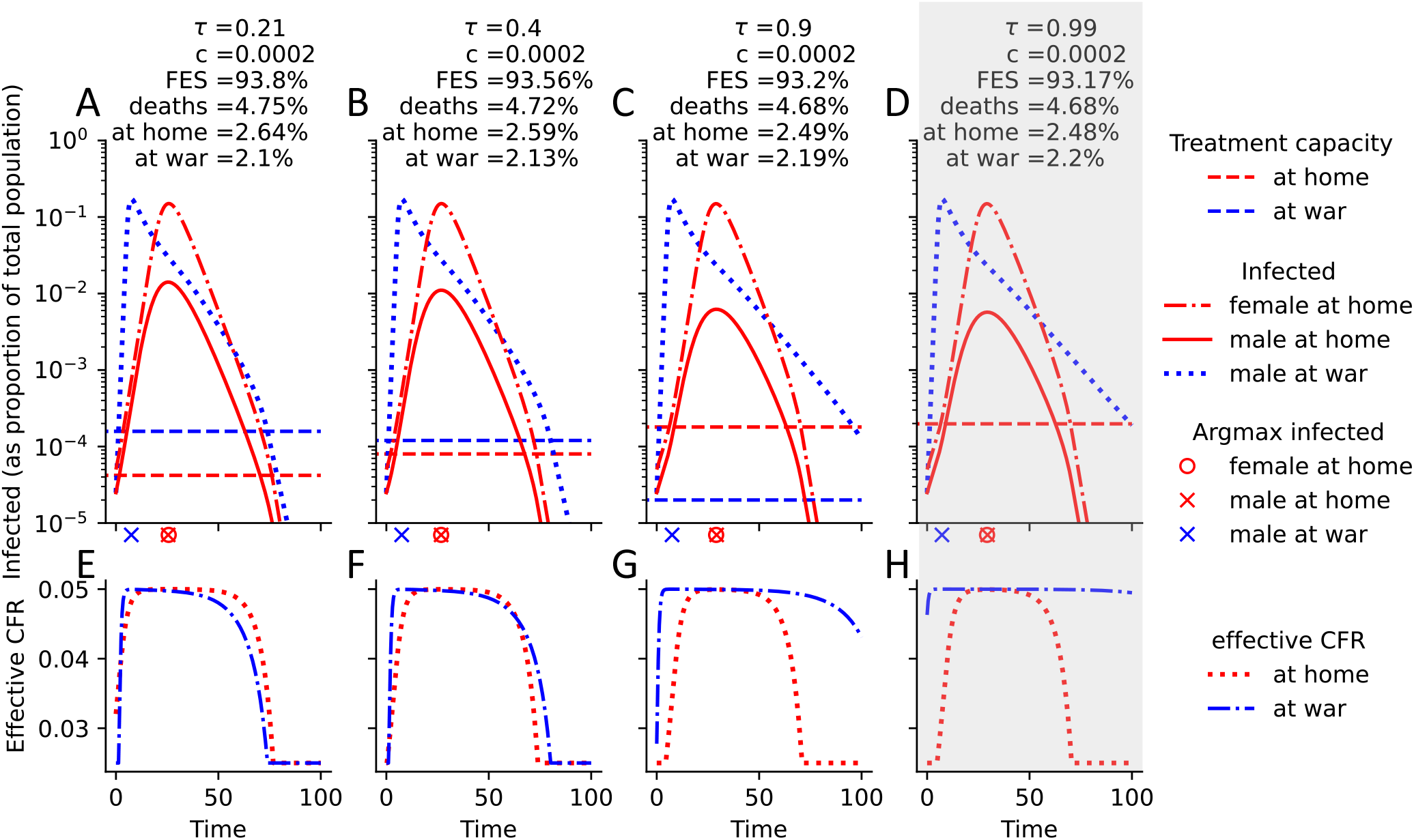
Comparison of epidemic curves when transmission rates at war are higher, males and females are completely segregated, and *c* = 0.0002 treatment resources are available. (A-D) For four different allocation strategies (A: *τ* = 0.21, B: *τ* = 0.4, C: *τ* = 0.9, D: *τ* = 0.99), the numbers of infected are shown over time, stratified by gender and location. For each choice of *τ*, the total resource capacity is fixed at *c* = 0.05, and the location-specific, *τ*-dependent resource availabilities are indicated by dashed lines. The location-specific epidemic peak is indicated just below the x-axis. Final epidemic size (FES), total deaths, deaths at home and at war are shown at the top. A gray box highlights the total mortality-minimizing strategy (D). All parameters are at their default value, as specified in Table 1, except for *β*_*W*_ = *β*_*H*_ = 0.6. (E-H) Corresponding plots of the effective case fatality rate (CFR), which is a function of the location-specific number of infected and treatment capacity, both shown in A-D.

**Figure S3:**
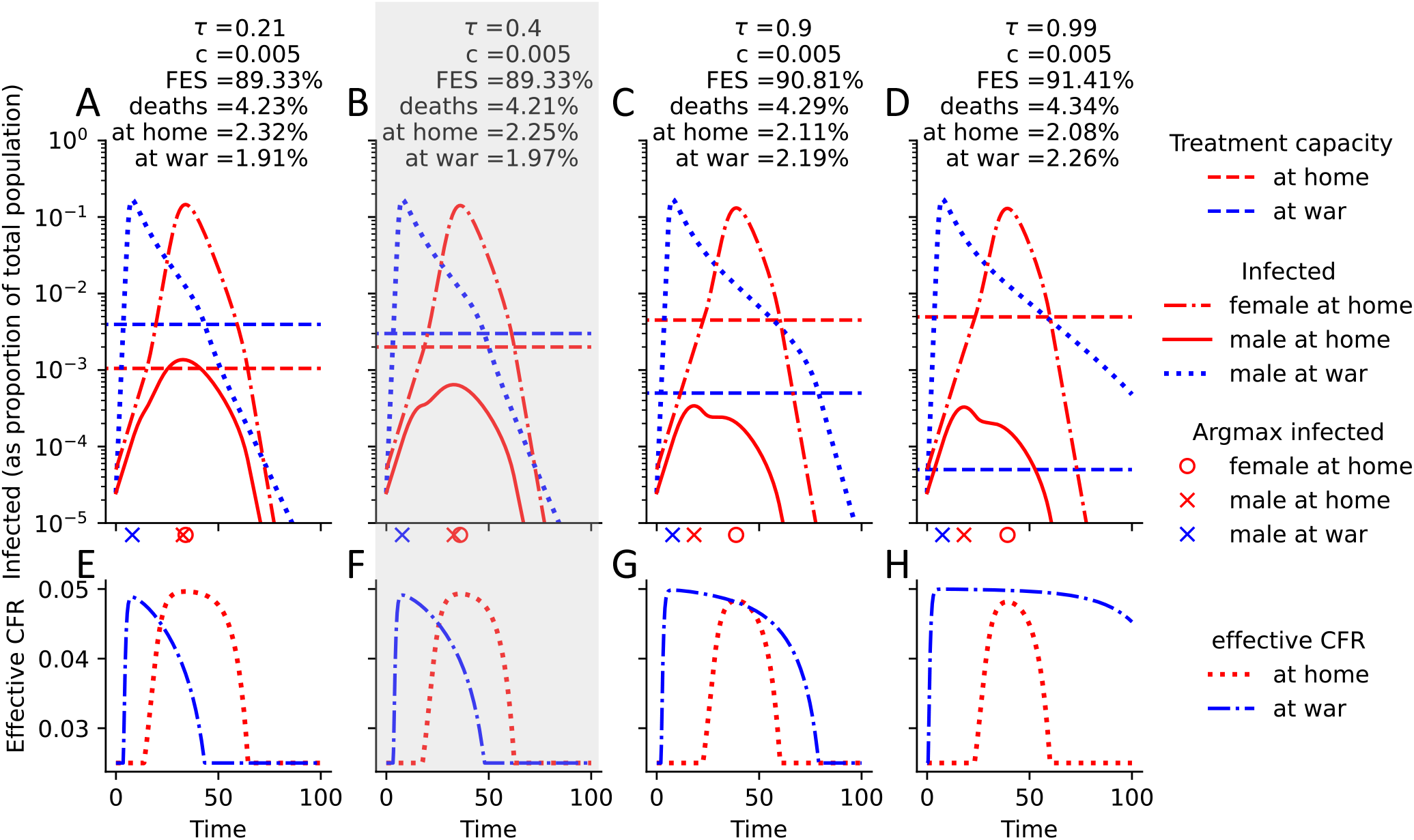
Comparison of epidemic curves when transmission rates at war are higher, males and females are completely segregated, and *c* = 0.005 treatment resources are available. (A-D) For four different allocation strategies (A: *τ* = 0.21, B: *τ* = 0.4, C: *τ* = 0.9, D: *τ* = 0.99), the numbers of infected are shown over time, stratified by gender and location. For each choice of *τ*, the total resource capacity is fixed at *c* = 0.05, and the location-specific, *τ*-dependent resource availabilities are indicated by dashed lines. The location-specific epidemic peak is indicated just below the x-axis. Final epidemic size (FES), total deaths, deaths at home and at war are shown at the top. A gray box highlights the total mortality-minimizing strategy (B). All parameters are at their default value, as specified in Table 1, except for *β*_*W*_ = *β*_*H*_ = 0.6. (E-H) Corresponding plots of the effective case fatality rate (CFR), which is a function of the location-specific number of infected and treatment capacity, both shown in A-D.

**Figure S4:**
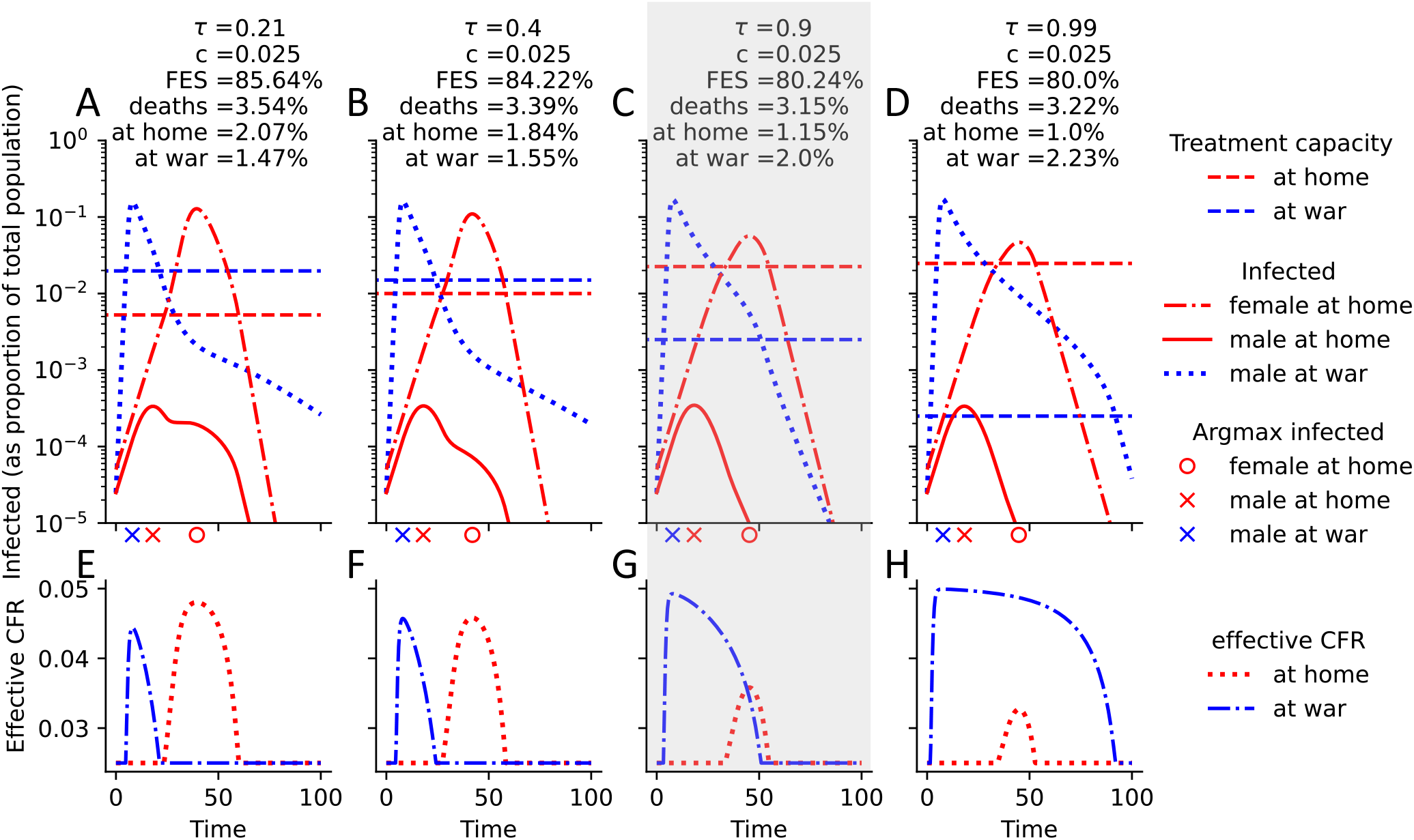
Comparison of epidemic curves when transmission rates at war are higher, males and females are completely segregated, and *c* = 0.025 treatment resources are available. (A-D) For four different allocation strategies (A: *τ* = 0.21, B: *τ* = 0.4, C: *τ* = 0.9, D: *τ* = 0.99), the numbers of infected are shown over time, stratified by gender and location. For each choice of *τ*, the total resource capacity is fixed at *c* = 0.05, and the location-specific, *τ*-dependent resource availabilities are indicated by dashed lines. The location-specific epidemic peak is indicated just below the x-axis. Final epidemic size (FES), total deaths, deaths at home and at war are shown at the top. A gray box highlights the total mortality-minimizing strategy (C). All parameters are at their default value, as specified in Table 1, except for *β*_*W*_ = *β*_*H*_ = 0.6. (E-H) Corresponding plots of the effective case fatality rate (CFR), which is a function of the location-specific number of infected and treatment capacity, both shown in A-D.

**Figure S5:**
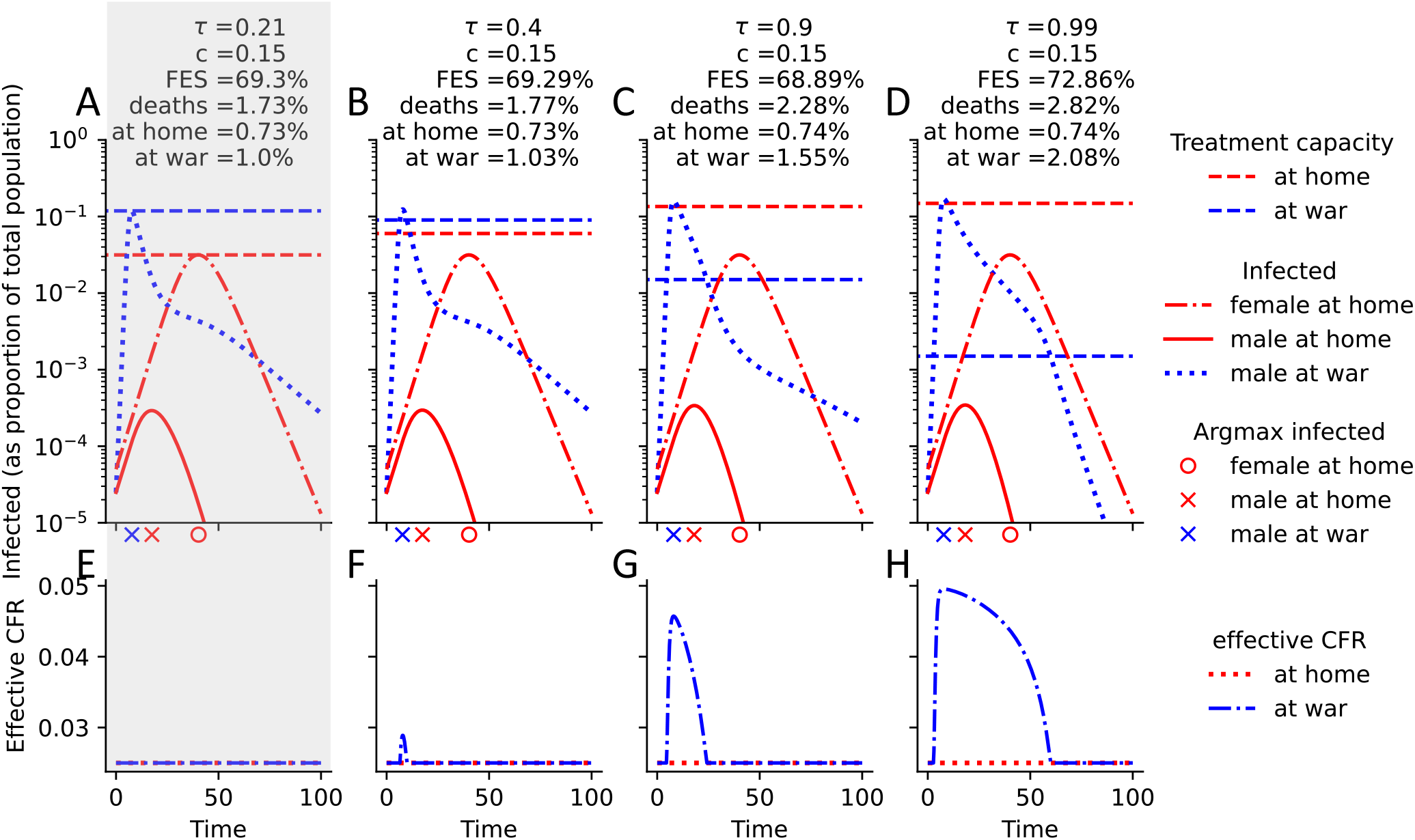
Comparison of epidemic curves when transmission rates at war are higher, males and females are completely segregated, and *c* = 0.15 treatment resources are available. (A-D) For four different allocation strategies (A: *τ* = 0.21, B: *τ* = 0.4, C: *τ* = 0.9, D: *τ* = 0.99), the numbers of infected are shown over time, stratified by gender and location. For each choice of *τ*, the total resource capacity is fixed at *c* = 0.05, and the location-specific, *τ*-dependent resource availabilities are indicated by dashed lines. The location-specific epidemic peak is indicated just below the x-axis. Final epidemic size (FES), total deaths, deaths at home and at war are shown at the top. A gray box highlights the total mortality-minimizing strategy (A). All parameters are at their default value, as specified in Table 1, except for *β*_*W*_ = *β*_*H*_ = 0.6. (E-H) Corresponding plots of the effective case fatality rate (CFR), which is a function of the location-specific number of infected and treatment capacity, both shown in A-D.

**Figure S6:**
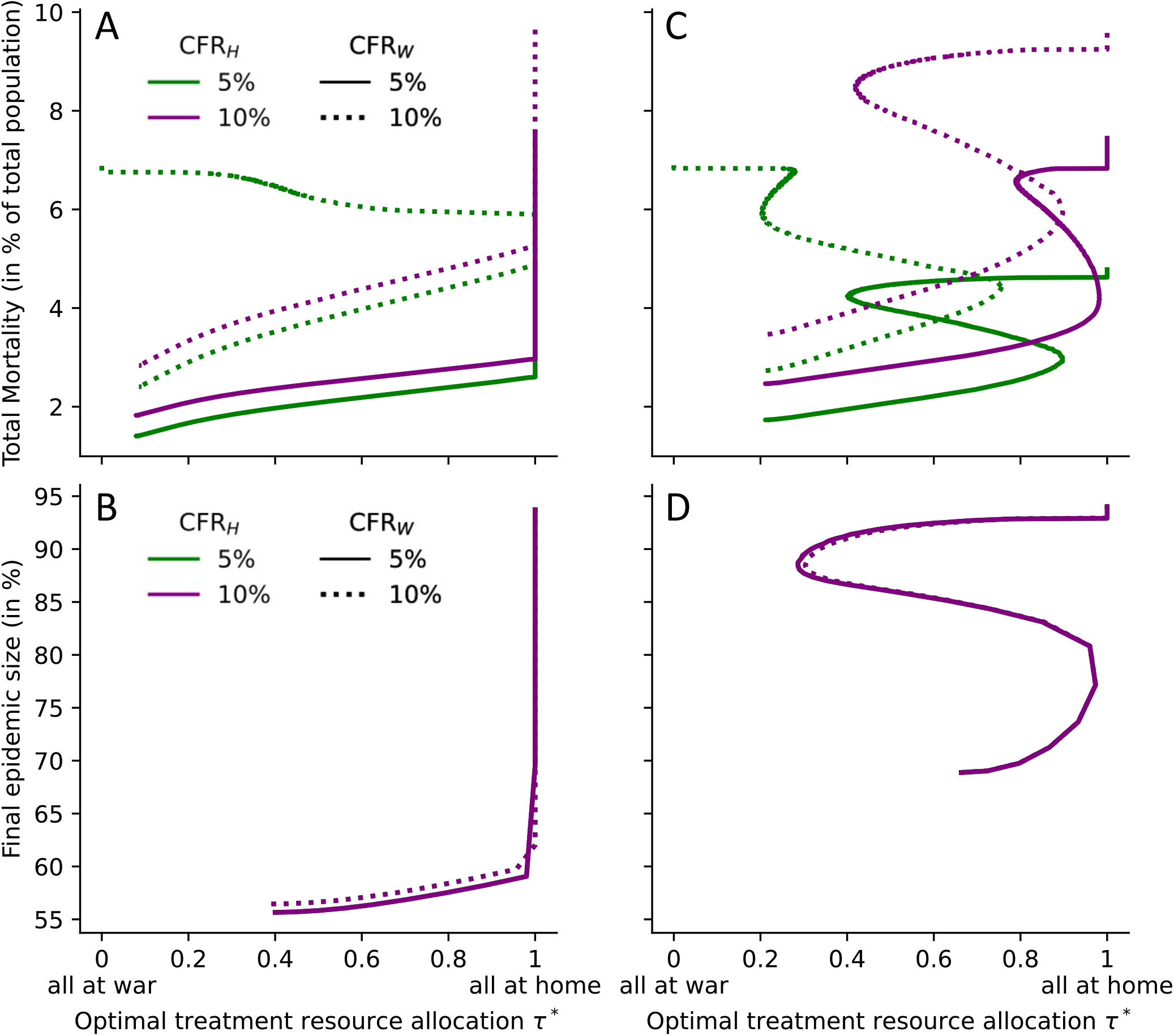
Optimal model-predicted allocation strategies for various treatment capacities and case fatality rates. The optimal allocation strategy *τ*^*^ is shown for any treatment capacity and two choices of the CFR at home and at war. The two extreme cases regarding gender homophily are considered: (A, B) no gender homophily (*h* = 0), (C, D) complete gender segregation (*h* = 1). Disease burden is quantified by (A, C) total mortality and (B, D) final epidemic size. All parameters are at their default value, as specified in Table 1, except for *β*_*W*_ = 3*β*_*H*_ = 1.8.

**Figure S7:**
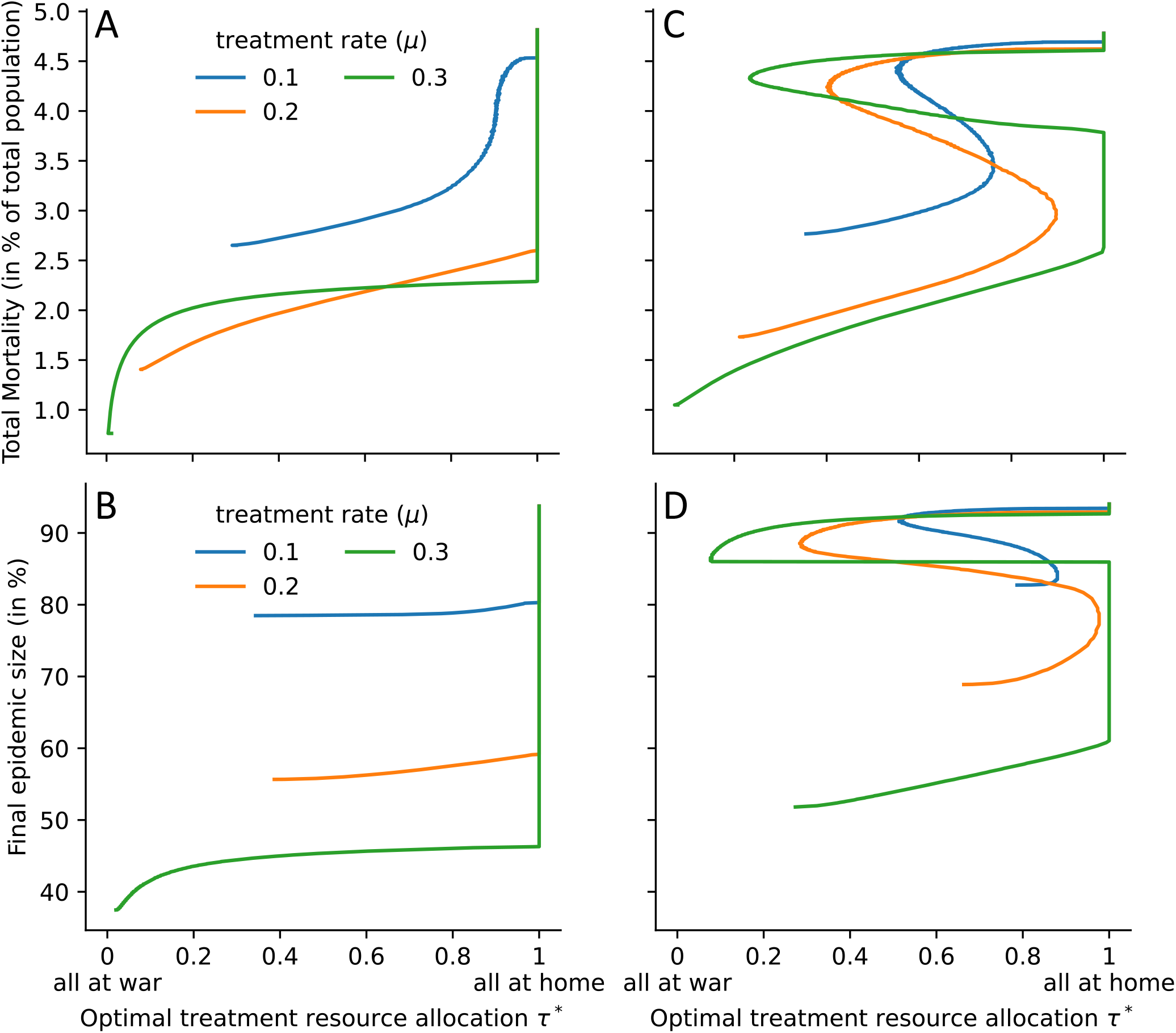
Optimal model-predicted allocation strategies for various treatment capacities and maximal treatment rates. The optimal allocation strategy *τ*^*^ is shown for any treatment capacity and three choices of maximal treatment rate. The two extreme cases regarding gender homophily are considered: (A, B) no gender homophily (*h* = 0), (C, D) complete gender segregation (*h* = 1). Disease burden is quantified by (A, C) total mortality and (B, D) final epidemic size. All parameters are at their default value, as specified in Table 1, except for *β*_*W*_ = 3*β*_*H*_ = 1.8.

**Figure S8:**
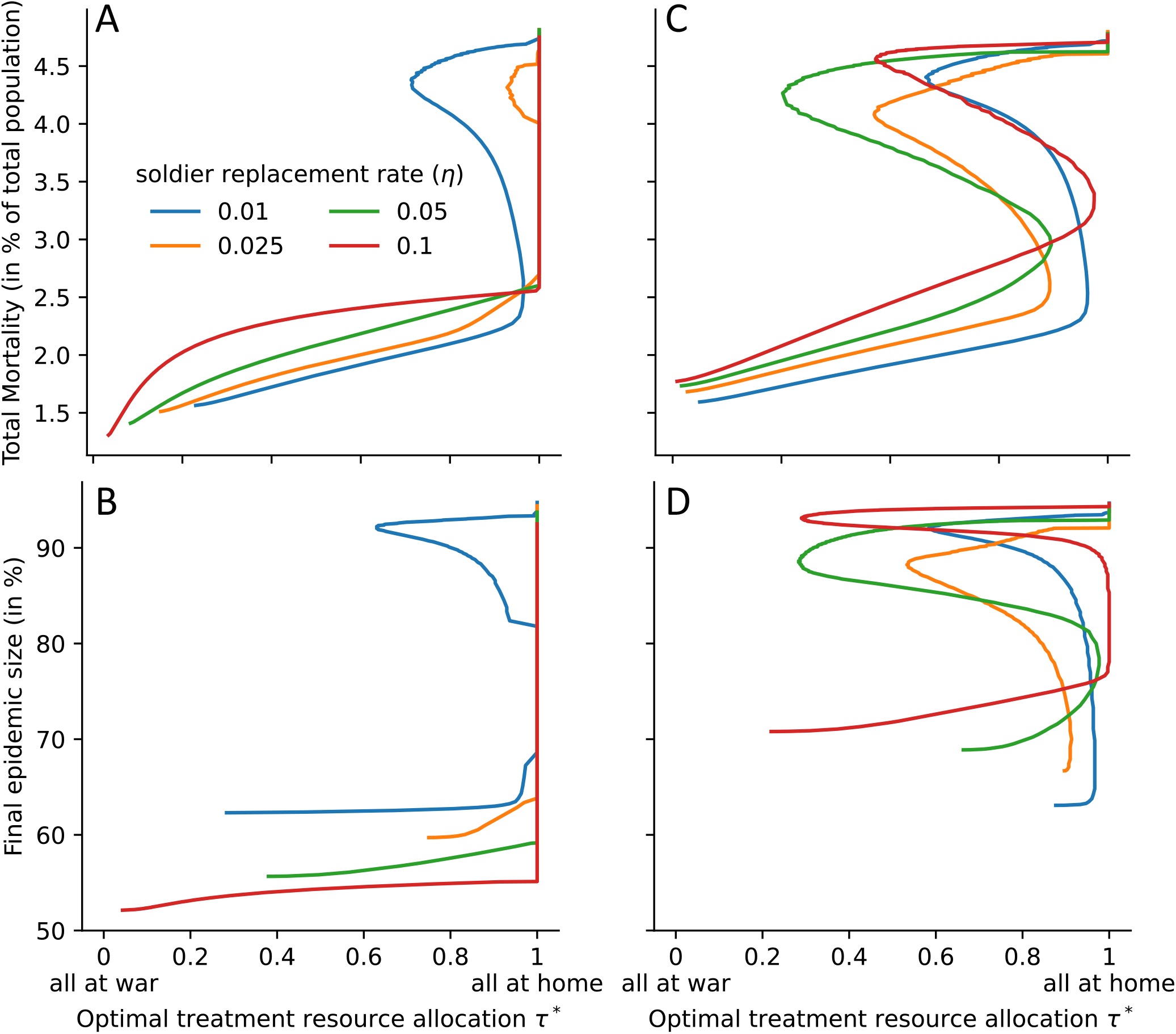
Optimal model-predicted allocation strategies for various treatment capacities and soldier replacement rates. The optimal allocation strategy *τ*^*^ is shown for any treatment capacity and three choices of maximal treatment rate. The two extreme cases regarding gender homophily are considered: (A, B) no gender homophily (*h* = 0), (C, D) complete gender segregation (*h* = 1). Disease burden is quantified by (A, C) total mortality and (B, D) final epidemic size. All parameters are at their default value, as specified in Table 1, except for *β*_*W*_ = 3*β*_*H*_ = 1.8.

**Figure S9:**
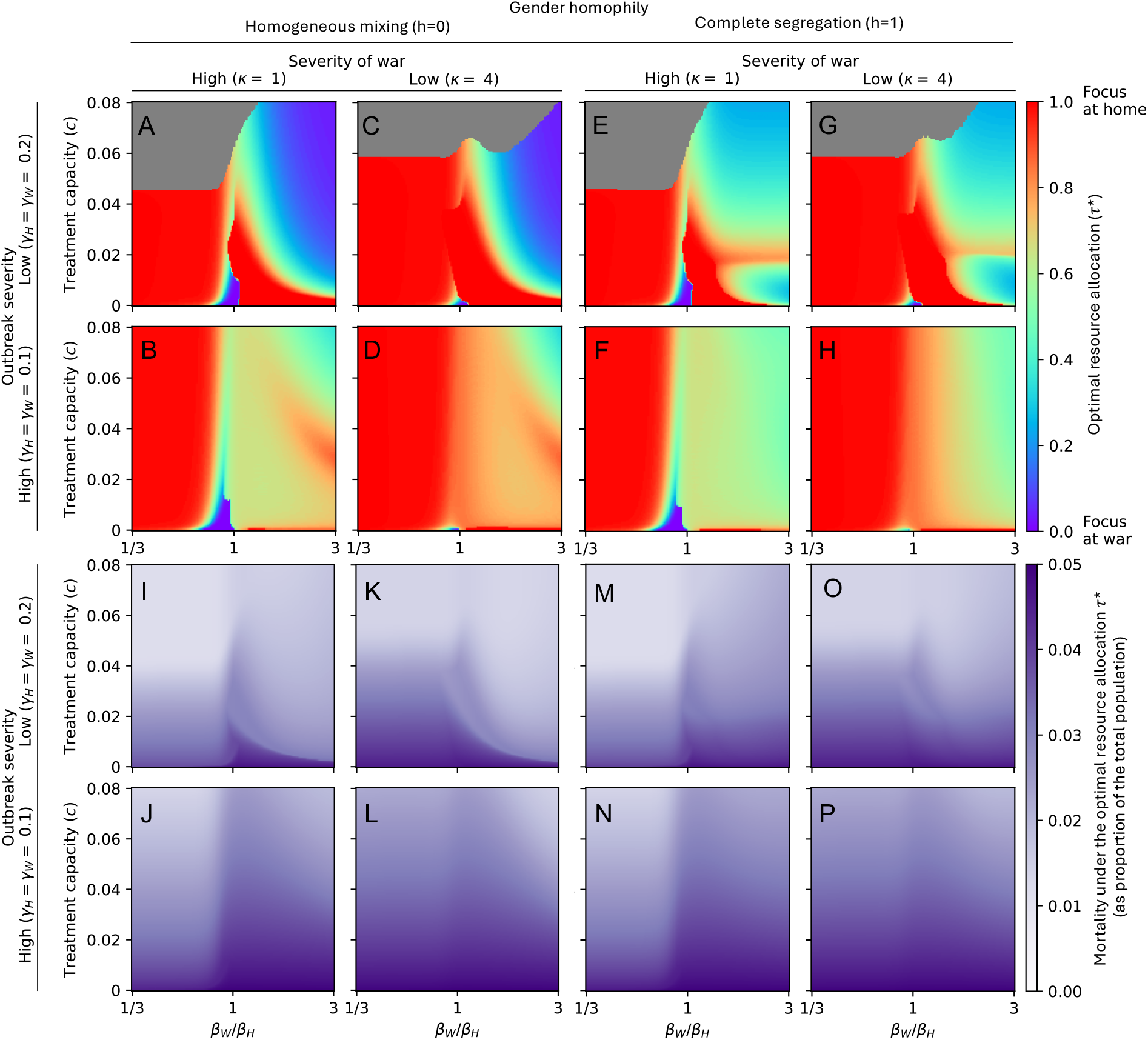
Sensitivity of the optimal resource allocation to changes in the male-to-female ratio. This figure shows the same five-dimensional sensitivity analysis as Fig. 5, except that 70% of the population is assumed to be male. For 150 equally-spaced values of treatment capacity (*c* ∈ [0, 0.08]) and transmission rates at war (*β*_*W*_ ∈ [1*/*3*β*_*H*_, 3*β*_*H*_]), the disease-induced mortality-minimizing treatment allocation strategy *τ*^*^ is shown in (A-H), while the total deaths under the respective strategy are shown in (I-P). The gray region describes parameter combinations where a range of resource allocations were optimal, i.e., where treatment resources sufficed to treat everyone optimally. In (A-D, I-L), males and females mix homogeneously (*h* = 0), while in (E-H, M-P), the extreme case of complete gender segregation (*h* = 1) is assumed. The outbreak severity varies between the rows, modulated by considering *γ*_*H*_ = *γ*_*W*_ = 0.2 (low severity) in rows 1 and 3, and *γ*_*H*_ = *γ*_*W*_ = 0.1 (high severity) in rows 2 and 4. The severity of war varies between the columns, modulated by considering *κ* = 1 (high severity, 50% of males at war) in rows 1 and 3, and *κ* = 4 (low severity, 20% of males at war) in rows 2 and 4. All other parameters are fixed at the default values described in Table 1.

**Figure S10:**
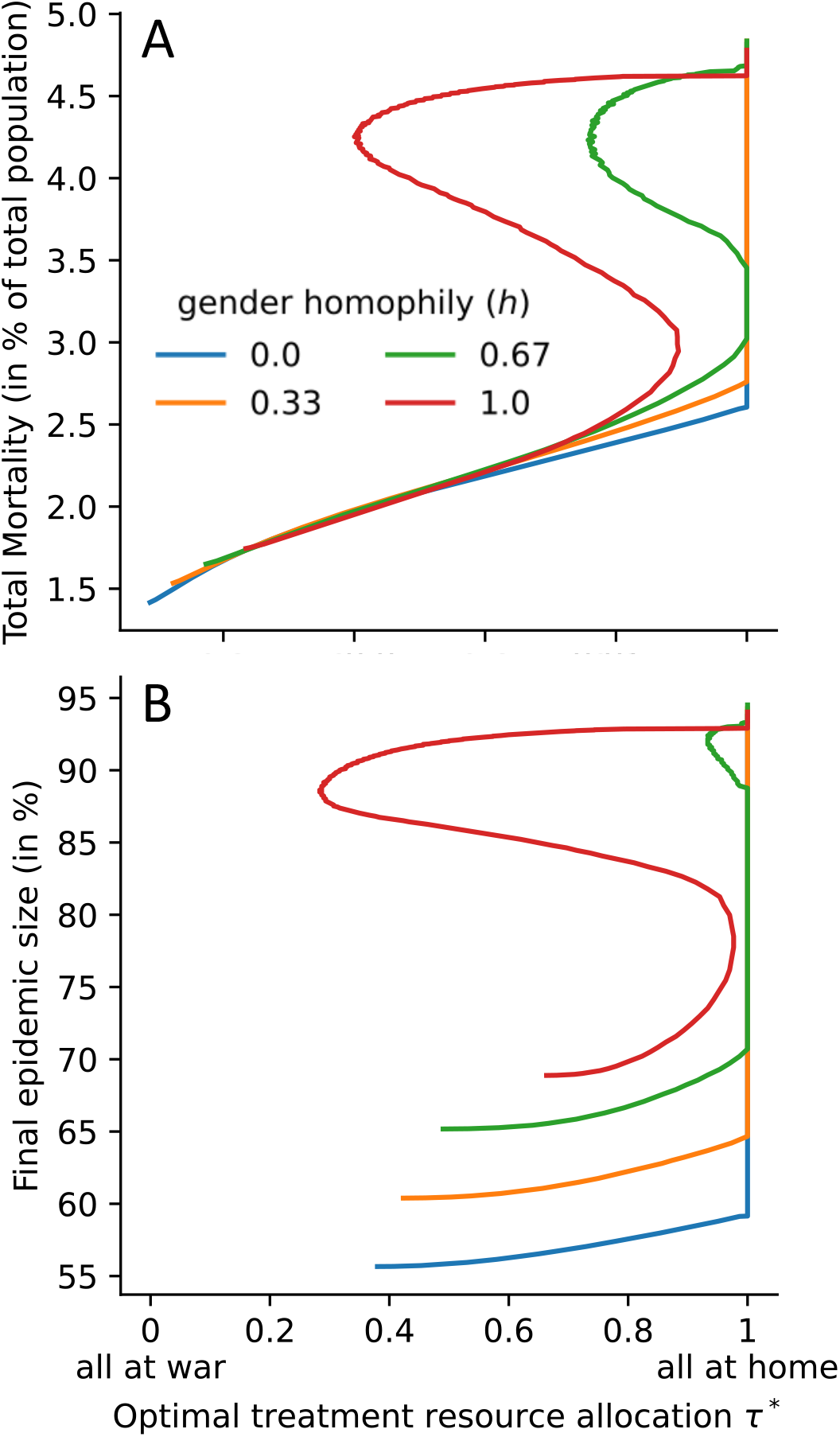
Optimal model-predicted allocation strategies for various treatment capacities and levels of gender homophily. The optimal allocation strategy *τ*^*^ is shown for any treatment capacity and three choices of maximal treatment rate. Disease burden is quantified by (A) total mortality and (B) final epidemic size. All parameters are at their default value, as specified in Table 1, except for *β*_*W*_ = 3*β*_*H*_ = 1.8.

## Notes

### Competing Interest Statement

The authors have declared no competing interest.

### Funding Statement

Vaibhava Srivastava gratefully acknowledges financial support from the Johnston-Peters Summer Graduate Assistantship. Claus Kadelka was partially supported by a travel grant from the Simons Foundation (grant number 712537).

